# Chronic health conditions and health-related economic inactivity in midlife: Evidence from the 1958 and 1970 British birth cohorts

**DOI:** 10.1101/2025.06.20.25329982

**Authors:** Laura Gimeno, Charis Bridger Staatz, Alice Goisis, Jennifer B. Dowd, George B. Ploubidis

## Abstract

**Background:** Health-related economic inactivity is a growing concern in the United Kingdom but little is known about how the relationship between health and work participation has changed across cohorts.

**Methods:** We used data from two British birth cohorts born in 1958 (National Child Development Study, *n* = 9,761) and 1970 (British Cohort Study, *n* = 7,336). We examined how self-reported chronic health conditions at age 42 (longstanding illness, obesity, diabetes, high blood pressure, back pain, and mental ill-health) were associated with economic activity at ages 50–54, focusing on health-related inactivity. Multinomial logistic regression models, adjusted for previous economic activity and sociodemographic characteristics, were used to estimate average marginal effects (AME).

**Results:** Poor health was more prevalent in the 1970c, including among those still working at age 50-54. Longstanding illness and mental ill-health were associated with a higher risk of health-related inactivity in both cohorts. A longstanding illness at age 42 was associated with a 6 percentage-point increase in health-related inactivity risk a decade later (AME_1958_ = 5.9 [95% Confidence Interval (CI) 2.7, 9.1], AME_1970_ = 6.5 [95%CI 3.4, 9.6]), and mental ill-health with a 4.5 percentage-point higher risk (AME_1958_ = 4.4 [95%CI 0.9, 7.9], AME_1970_ = 4.5 [95%CI 1.1, 7.8]). The magnitude of associations was similar across cohorts except for high blood pressure.

**Conclusions:** Chronic health conditions in early midlife were strongly associated with a health-related inactivity, despite contextual change. Preventing ill-health and supporting employment for those with chronic conditions is key to face the challenges of population ageing.

## INTRODUCTION

Health-related economic inactivity (i.e., not working and not seeking work due to health problems) is a growing concern in the United Kingdom (UK). The Labour Force Survey estimated that 2.83 million people aged 16-64 were inactive due to long-term health problems (including mental ill-health, cardiovascular disease and musculoskeletal problems) in February-April 2024, up from 2 million in spring 2019 (Office for National Statistics, 2022, 2023, 2024). While methodological concerns with the Labour Force Survey cast doubt over the true magnitude of the increase in health-related inactivity since the COVID-19 pandemic (Corlett, 2024; Office for National Statistics, 2025), there is some suggestion this may be part of a longer-term trend of rising health-related inactivity beginning before pre-pandemic (Office for National Statistics, 2022).

Rising health-related inactivity presents an important fiscal challenge as it is linked to increased benefits expenditure, lower revenue from income tax, and reductions in National Insurance contributions. Health-related inactivity also often represents a sudden loss of income affecting the long-term financial stability of the affected individual and of their financial dependents (Brand, 2015). While health-related inactivity may be inevitable for many individuals, 1-in-4 people inactive due to their health state that they would like to work (Tinson et al., 2022). Reducing health-related inactivity – particularly among older workers – by improving population health and by better supporting people with chronic conditions to stay in work, constitutes a potentially attractive opportunity to mitigate the impact of population ageing on labour supply (André et al., 2024).

Associations between health across the lifecourse and many social and economic outcomes, including employment status and stability have been studied using longitudinal data in the UK (Gondek et al., 2018). Studies have quantified associations between health and all-cause labour market exit, transitions into retirement and unemployment, and, to a lesser extent, with health-related benefit receipt (Blundell et al., 2023; Disney et al., 2006; Henderson et al., 2024; Jones et al., 2020; O’Donnell et al., 2015; Rice et al., 2011; Stafford et al., 2017; Trevisan & Zantomio, 2016), and have found that those with chronic health conditions are more likely to drop out of work. While many individuals who are inactive due to health problems are in receipt of benefits, the overlap is not complete. Disability benefits can be received by those in work, and among those inactive due to health in 2022/3, 63% received disability benefits and 83% incapacity benefit (Office for Budget Responsibility, 2023). Health-related inactivity is an interesting outcome, since it excludes those who work, even part-time, and captures individuals who are not working due to their health but who may not be in receipt of health-related benefits and would otherwise be assumed to be inactive for other reasons.

Research exploring the causal impact of health on work participation has focused on a narrow range of “health shocks” (first cancer diagnosis, heart attacks, strokes, road traffic accidents), and has shown that experiencing a deterioration in health results in a higher risk of labour market exit (Disney et al., 2006; Jones et al., 2020). While helpful for establishing a causal relationship between health economic activity, these exposures have limited generalisability since the burden of ill-health in the working-age population in the UK is primarily chronic in nature. Other studies have leveraged changes in self-rated general health (Henderson et al., 2013), which may be more generalisable, but do not relate the risk of exits from the labour force to particular health conditions. Studies have explored the impact of mental ill-health, chronic pain, musculoskeletal problems on work participation in the UK, but these studies have focused on transitions to early retirement rather than health-related inactivity (Rice et al., 2011; Stafford et al., 2017).

The extent to which increases in health-related inactivity and benefits claims are due to long-term worsening population health is an area of active debate (Atwell et al., 2023; Boileau & Cribb, 2022; Cribb, 2023). Increases in the prevalence of obesity and mental ill-health have been observed across cohorts (Armitage et al., 2023; Gondek et al., 2018; Johnson et al., 2015; McElroy et al., 2023). However, associations between health and health-related inactivity could also have changed over time, such that someone with a chronic health condition has a higher (or lower) risk of being inactive due to health today than they would have done in the past. Between 1973 and 2009, economic inactivity rates among those in poor health increased among men, and remained stable among women despite declining in women in good health (Minton et al., 2012). However, these data are cross-sectional, and the impact of reverse causality cannot be ruled out. There have also been policy changes increasing the conditionality of health-related benefit receipt, and laws against discrimination on the grounds of disability have been introduced (Figure S1). Such policies, coupled with a shift to non-manual occupations in more recently born cohorts, could have resulted in individuals with chronic health conditions being more likely to remain economically active than they might have been in the past. Examining whether associations between health and health-related inactivity differ across cohorts could shed light on this debate.

In this study, we used data from two British birth cohort studies of people born 12 years apart (in 1958 and 1970) to explore associations between chronic health conditions in early midlife and economic activity in cohort members’ early fifties, more than a decade before State Pension Age. We describe changes in health by economic activity status across cohorts and quantify associations between health and economic activity in each cohort.

## METHODS

### Data

We used data from the 1958 National Child Development Study (1958c) and the 1970 British Cohort Study (1970c). These surveys have followed all people born in England, Wales and Scotland in one week of 1958 (*n* = 18,558) and 1970 (*n =* 18,037) from birth until today. The history and design of these studies has been described in detail elsewhere (Elliott & Shepherd, 2006; Power & Elliott, 2006; Sullivan et al., 2023). Ethical approval for both studies was received from the NHS Multi-Centre Research Ethics Committee, with informed consent provided by all participants. We used data collected up to age 50-51 in the 1958c (collected 2008-2009), and age 51-54 in the 1970c (collected 2021-2024). While age overlap was not exact, these data collection sweeps correspond to the first surveys in cohort members’ fifties in each cohort study.

Το descriptive the distribution of health and economic activity at age 42, we used data from all cohort members who responded to the age 42 sweeps in the 1958c (*n =* 11,419) and 1970c (*n =* 9841). Subsequent analyses exploring the relationship between health at age 42 and economic activity in cohort members’ fifties used data from all respondents to the age 50-51 sweep of the 1958c (*n =* 9841) and the age 51-54 sweep of the 1970c (*n =* 7336) with information on current economic activity (Figure 1).

**Figure 1.**
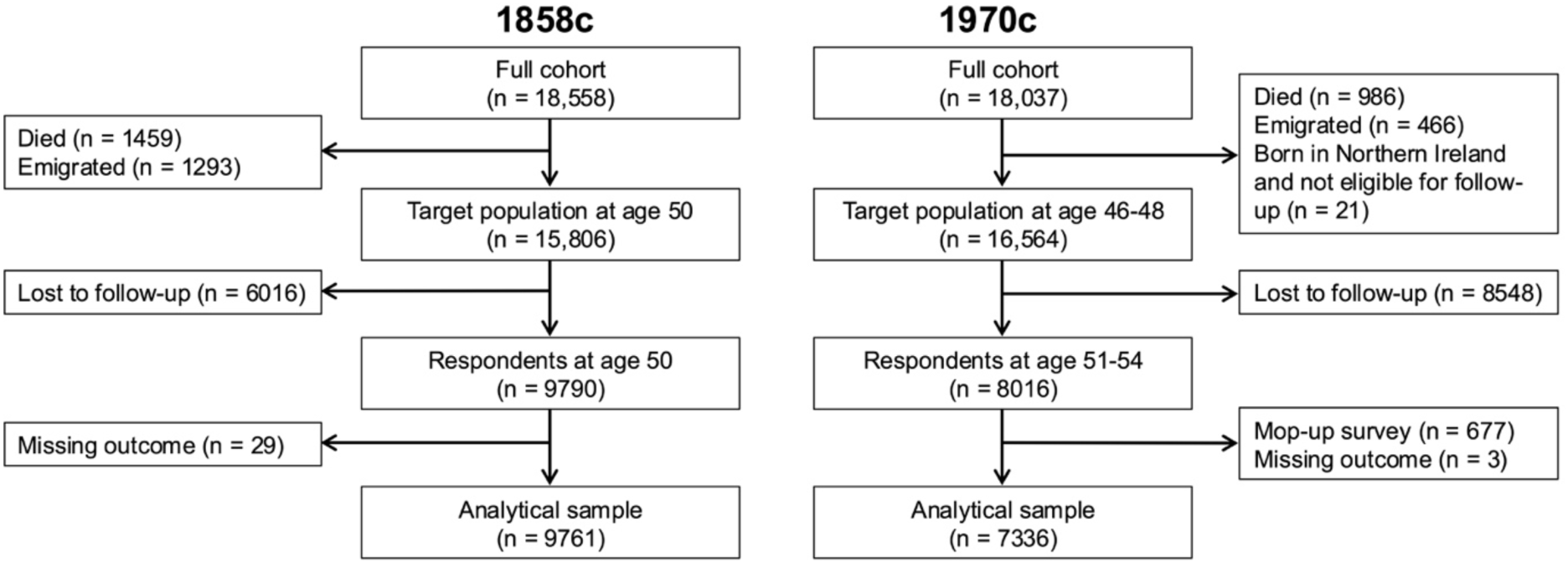
**Derivation of the analytical sample in the 1958c and 1970c**.

### Outcome

The outcome variable was economic activity in cohort members’ fifties, grouped into five categories:

- Active full-time (ACT-FT): employed or self-employed working >30 hours per week
- Active part-time (ACT-PT): employed or self-employed working ≤ 30 hours per week
- Unemployed and looking for work (UNEMP)
- Inactive due to health (IN-HLT): Includes those inactive due to both long-term and short-term sickness
- Inactive due to other reasons (IN-OTH). Reasons include full-time education, training schemes, looking after home or family, retired, other.

We briefly report findings for all outcome levels but focus specifically on IN-HLT, given the current lack of longitudinal evidence for this outcome.

### Exposures

Our analyses focused on self-reported health conditions which reflect major causes of health-related economic inactivity identified in the Labour Force Survey: poor mental health, cardiometabolic disease, and musculoskeletal disease. More details on the exposures are given in Table S1.

- ***Longstanding illness***. Whether cohort members reported any longstanding illness or disability at age 42. This single question about general health is widely used in social surveys and in the census and has been shown to correlate strongly with reports of specific, more severe health conditions (Ayis et al., 2003; Manor et al., 2001).
- ***Psychological distress***. Whether cohort members scored ≥4 on the 9-item version of the Malaise Inventory (Rutter et al., 1970), which measures non-specific symptoms of depression, anxiety and distress.
- ***Obesity***. Whether cohort members had a body mass index ≥30 kg/m^2^ at age 42.
- ***Diabetes***. Whether cohort members had ever had or been told they had diabetes up to and including age 42.
- ***High blood pressure***. Whether cohort members had ever had or been told they had high blood pressure up to and including age 42.
- ***Back pain***. Whether cohort members had ever had or been told they had persistent back pain, sciatica, or slipped disc up to and including age 42.

### Confounders

A range of socioeconomic and demographic variables associated with both health status and economic activity were included in the analysis (Table 1).

**Table 1.** Covariates included regression models.

| Variable | Definition | Age measured |  |
| --- | --- | --- | --- |
|  |  | NCDS | BCS70 |
| Economic activity (lagged outcome) | Active full-time, active part-time, unemployed, inactive due to health, inactive other causes | 42 | 42 |
| Sex at birth | Male, female | 0 | 0 |
| Father's social class | Professional/managerial/intermediate/skilled (RGSC I-III), partly skilled/unskilled (RGSC IV-V) | 0 | 0 |
| Cognitive ability | Principal component 1 from PCA of cognitive test scores | 11 | 10 |
| Region of residence | North/North West/Yorkshire and Humberside, West Midlands/South West, East Midlands/East Anglia, South East/London, Wales/Scotland/Northern Ireland | 42 | 42 |
| Partnership status | Married/cohabiting, other | 42 | 42 |
| Educational attainment | No degree (NVQ levels 0-3), degree (NVQ level $\geq 4$) | 42 | 42 |
| Equivalised household income | Quintile of equivalised household income. Includes income from cohort member, their partner, and any additional sources of income (e.g., benefits), and is related back to the size of the household. | 42 | 42 |
| Housing tenure | Owns home or is buying home with the help of a mortgage, other. | 42 | 42 |
| Children in household | Whether there are any (biological or non-biological) children resident in the household. | 42 | 42 |
| Occupation | Non-manual (RGSC I-IIINM), non-manual (IIIM-V) | 42 | 42 |
| Age at outcome measurement | Age when the outcome was measured in years (year surveyed minus year of birth) for BCS70 only | NA | 51-54 |

**Note:** RGSC = Registrar General Social Class 1990. PCA = Principal Component Analysis. NVQ = National Vocational Qualification. Patterns of missing data are described in Supplementary Text 1.

### Descriptive Statistics

We described the prevalence of poor health at age 42 and the distribution of economic activity in in both cohorts. We then examined the prevalence of poor health within economic activity categories, and how cohort members with health conditions at age 42 were distributed across economic categories at age 50-54 to quantify cohort changes in health amongst those who remained in the labour market. We also described proportion of cohort members who were inactive due to health problems at age 50-54 who were already in this category at age 42.

### Main Analysis

Using multinomial regression models stratified by cohort, we examined how health status at age 42 was associated with economic activity a decade later. We focused on health-related economic activity as the primary outcome of interest, though we report results for other economic activity categories.

Unadjusted models included only economic activity at age 50-54 as the dependent variable (with full-time activity as the reference category) and each health condition at age 42 in separate models as the exposure. In fully adjusted models, we additionally included a lagged a lagged outcome (economic activity at age 42), and demographic and socioeconomic confounders from across the lifecourse including early-life factors such as cognitive ability, parental social class, and sex at birth (Table 1). The lagged outcome was included to (i) account for the bi-directional relationship between health and employment, recognising that individuals with health conditions were more likely to be economically inactive at baseline; and, crucially, (ii) to help block pathways from unmeasured confounders to the outcome, as illustrated in Figure 2, thereby strengthening causal inference. The inclusion of a broad set of life course confounders in the fully adjusted model also enhances the plausibility that the adjusted associations reflect causal effects, by reducing the risk of residual confounding.

**Figure 2.**
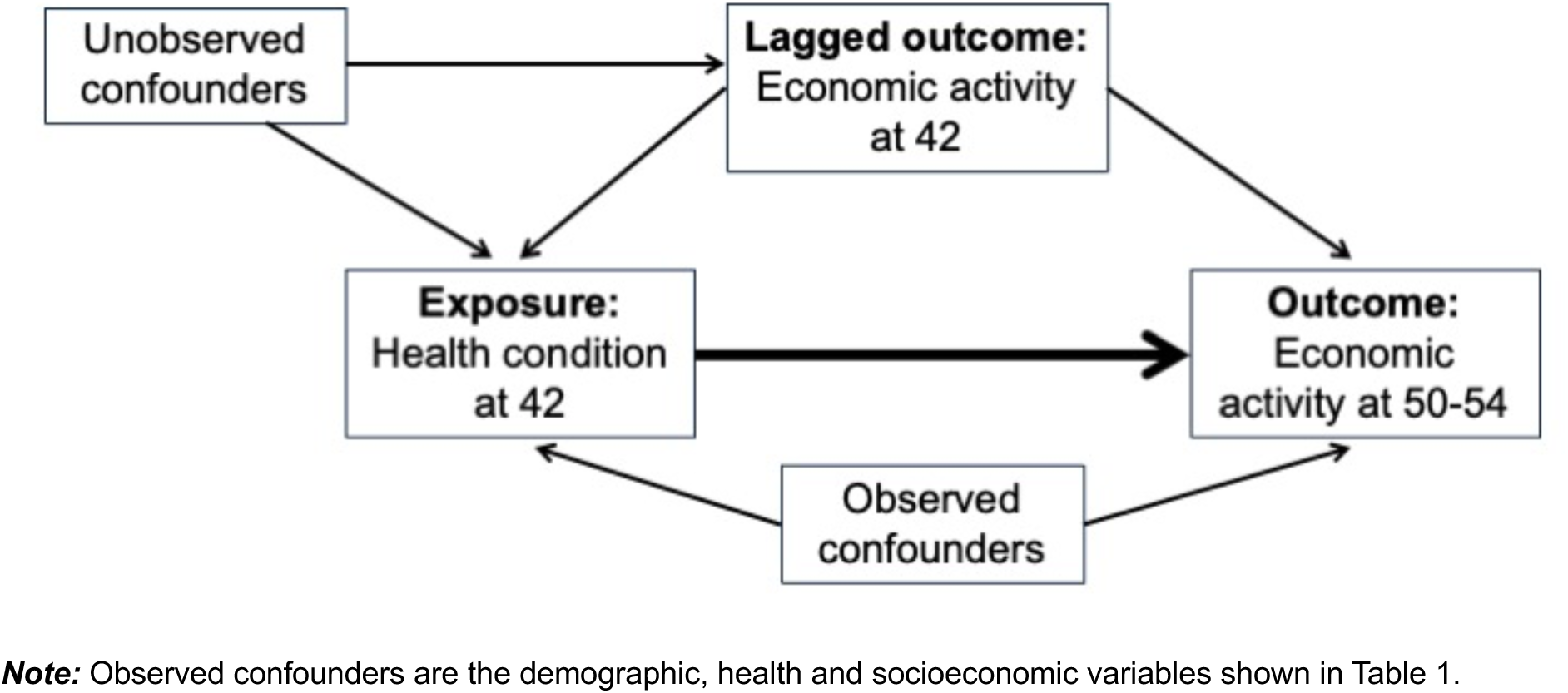
**Directed Acyclic Graph showing proposed relationship between variables included in the main analysis**.

To facilitate interpretation, we report results as average marginal effects (alongside the predicted risk of the outcome among those without the health condition). Average marginal effects estimate the percentage point change in the risk of being in each economic activity group (e.g., the difference in the risk of being in the IN-HLT group at age 50-54 for individuals with and without longstanding illness at age 42), accounting for differences in previous employment status and life course sociodemographic characteristics.

### Secondary Analysis

We described the gender and socioeconomic (educational attainment, housing tenure, occupation) composition of two groups within and across birth cohorts: those with poor health at age 42 in the ACT-FT group at age 50-54, and those with poor health in the IN-HLT group at age 50-54.

### Sensitivity Analysis

We carried out a series of sensitivity analyses, described in more detail in the Supplementary Material. These included (1) replicating the analysis when ACT-FT, ACT-PT and UNEMP were grouped into a single economically active category; (2) additionally adjusting for early-life potential confounders (maternal and paternal education, housing tenure at age 5/7, breastfeeding, and maternal smoking); (3a) restricting to those ACT-FT, ACT-PT or UNEMP at age 42 to remove the impact of reclassifications across economic inactivity categories, and (3b) excluding those IN-HLT at age 42.

We also explored whether the direction of associations was similar across subgroups (by gender, educational attainment, household income quintile, housing tenure, occupation) for two health conditions: longstanding illness and psychological distress.

### Missing Data Strategy

Like all longitudinal studies, the 1958c and 1970c have experienced loss to follow-up, the likelihood of which varies by individual level characteristics, such as gender, health, and socioeconomic status. To mitigate the impact of loss to follow-up, we capitalised on the richness of the cohort data and the known properties of the cohort members, handling item missingness using multiple imputation by chained equations (MICE), and using inverse-probability weights to handle sweep non-response (Katsoulis et al., 2024; Mostafa et al., 2021). More details on the missing data strategy can be found in the Supplementary Material.

All analyses were conducted using Stata version 18.0 (StataCorp, College Station, TX).

## RESULTS

### Descriptive Analysis

The prevalence of almost all health conditions at age 42 was higher in the 1970c compared to the 1958c, except for longstanding illness (Figure 3, Table S2). For instance, those in the 1970c were more likely to be obese (23.9% vs. 16.8%), to be experiencing psychological distress (21.4% vs. 14.2%) and to have self-reported histories of back pain (31.0% vs. 23.1%), high blood pressure (13.5% vs. 11.9%), or diabetes (3.3% vs. 2.6%, 95% confidence intervals overlap).

**Figure 3.**
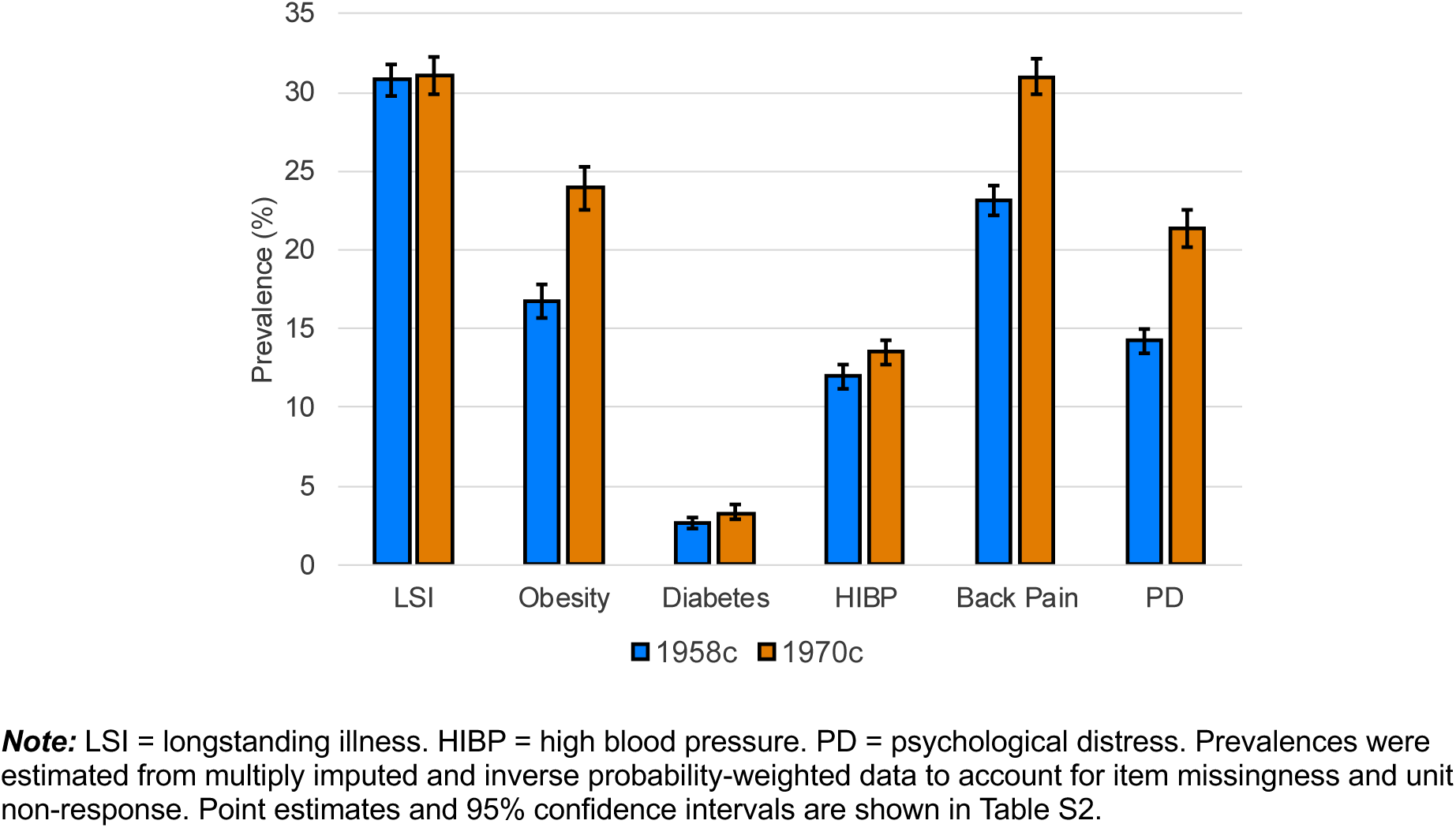
**Prevalence of poor health at age 42**.

The prevalence of IN-HLT in each cohort was higher in cohort members’ early fifties than at age 42 (Figure 4, Table S3). The distribution of economic activity was similar across cohorts, though the proportion unemployed and inactive was slightly higher in the 1958c at age 50-51 (in 2008/9) and in the 1970c at age 42 (in 2012), highlighting the impact of the 2008 Great Recession on these cohorts at different ages (Figure S2). The prevalence of IN-OTH was also lower in the 1970c, likely due to higher labour force participation among women in this cohort. The proportion of cohort members who were IN-HLT at age 50-54 was similar across cohorts: 8.5% (95% CI 7.5-9.4) in the 1958c, and 8.8% (95% CI 7.4-10.2) in the 1970c. In both cohorts, approximately half of cohort members who were IN-HLT at age 50-54 were also IN-HLT at age 42 (Table S4-S5).

**Figure 4.**
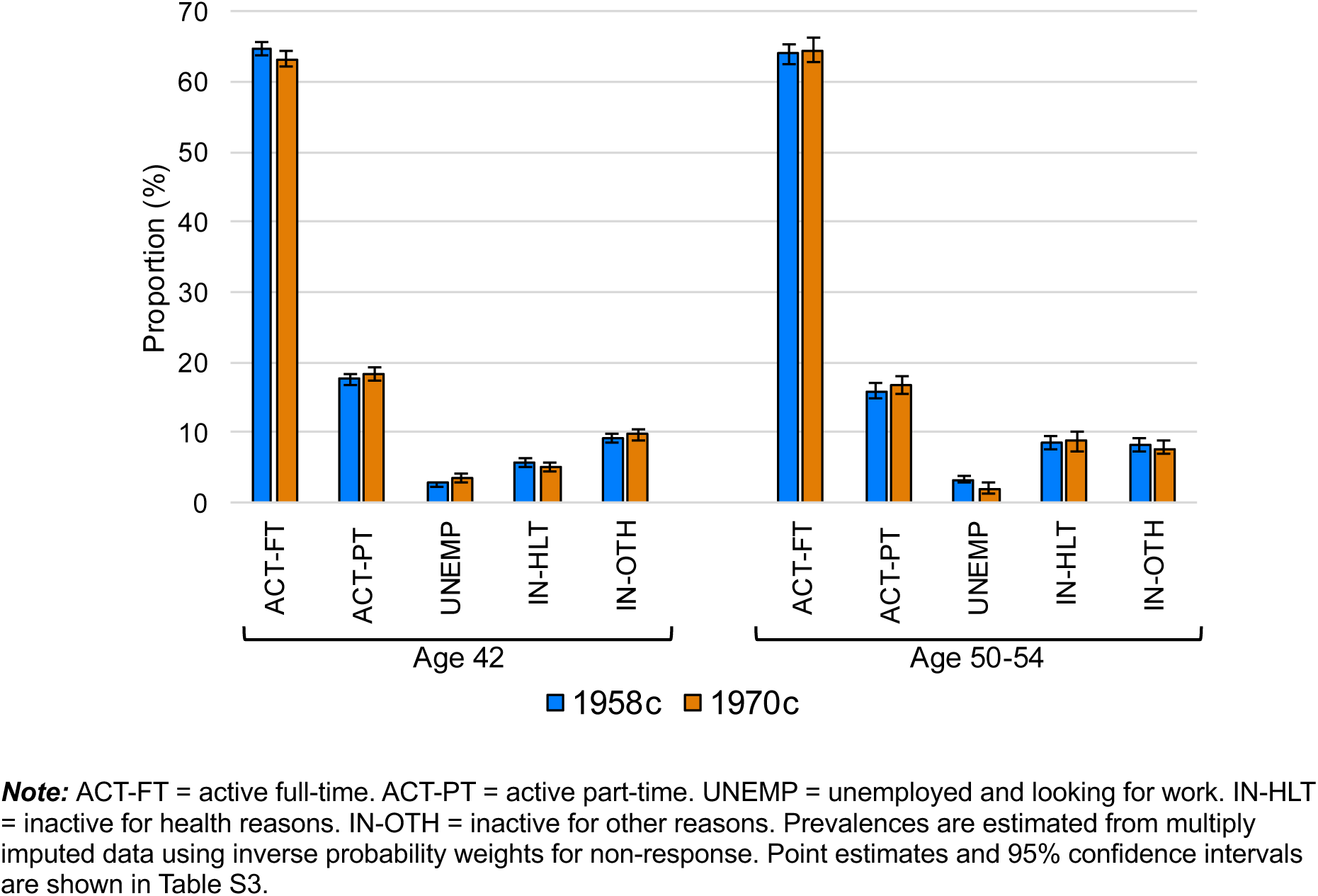
**Distribution of economic activity at ages 42 and 50-54 in the 1958c and 1970c**.

The prevalence of obesity, psychological distress, and back pain among those ACT-FT at age 50-54 was higher in the 1970c (Table S6, Figure S3), generally reflecting the increase in prevalence of long-term health conditions across cohorts. However, this prevalence increase was also observed in the IN-HLT group, except for high blood pressure (Table S6, Figure S3). The proportion of cohort members with chronic conditions at age 42 who were working (ACT-FT and ACT-PT) at age 50-54 was generally similar or higher in the 1970c than the 1958c, though 95% confidence intervals overlapped (Figure S4). However, for several conditions, the proportion of cohort members with chronic health conditions who were IN-HLT at age 50-54 also increased across cohorts, while the proportion unemployed or IN-OTH declined.

### Main Analysis

We quantified associations between health conditions at age 42 and economic activity at age 50-54. While ACT-FT, ACT-PT, UNEMP, IN-HLT and IN-OTH are each outcomes in their own right, we focus our interpretation on the IN-HLT group. Results for other outcomes are briefly summarised, and results are shown in full in the Supplementary Material.

In unadjusted models, without accounting for differences in previous economic activity or sociodemographic characteristics, those with a chronic health condition were significantly more likely to be IN-HLT in their early fifties in both cohorts compared to those without the condition (Figure 5).

**Figure 5.**
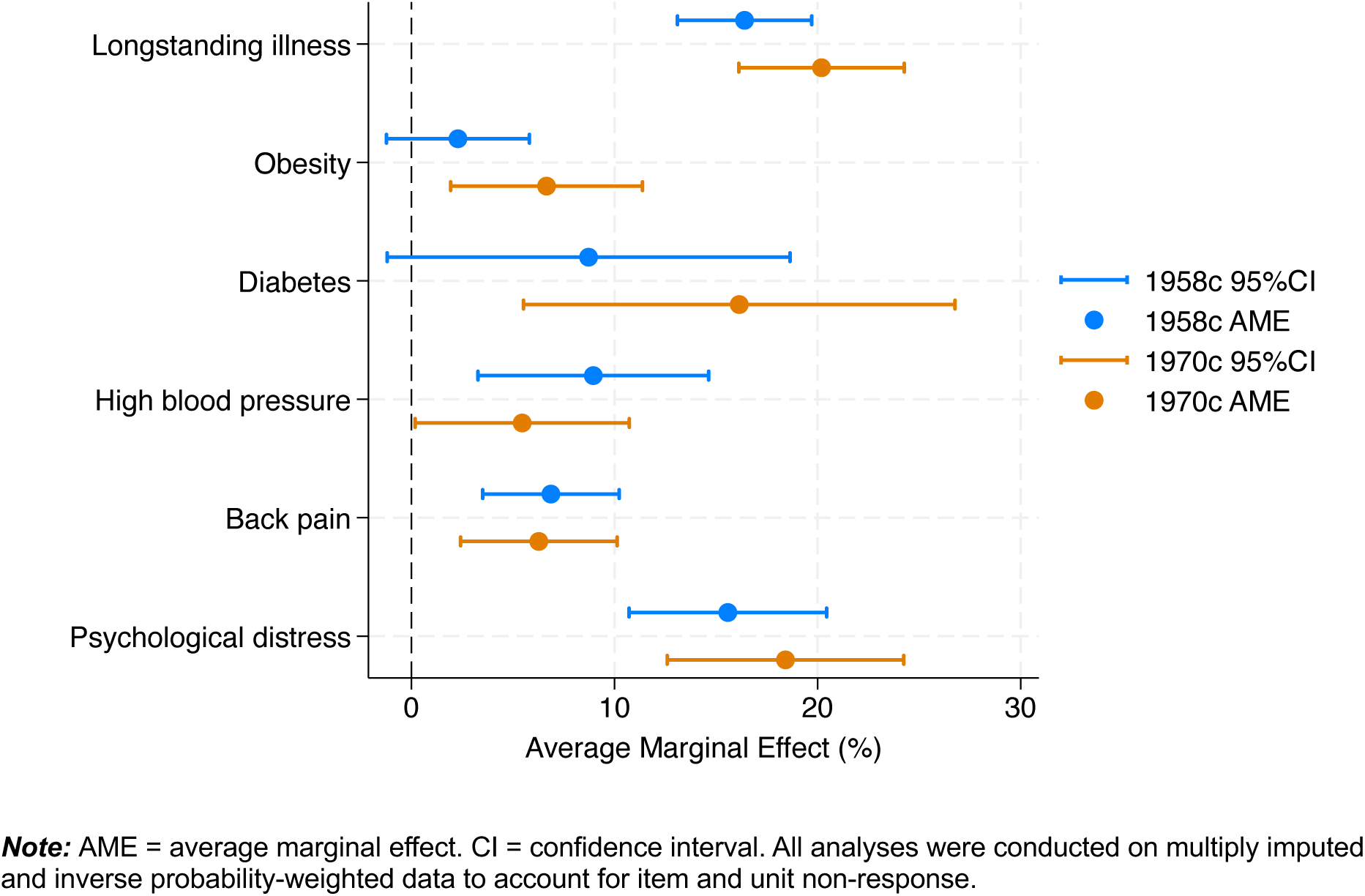
**Average marginal effects from unadjusted models**.

For instance, having a longstanding illness at age 42 was associated with a 16-20 percentage-point higher risk of being IN-HLT at age 50-54 (Average Marginal Effect (AME)_1958_ = 16.4 [95%CI 13.1, 19.7], AME_1970_ = 20.2 [95%CI 16.1, 24.3]). Predicted probabilities of IN-HLT for those with and without health conditions were relatively similar across cohorts. While among those without each health condition the predicted probability of IN-HLT tended to be the same or lower in the 1970c compared to the 1958c, for most conditions, there was some suggestion that the predicted probability of IN-HLT was higher in the 1970c for those with several chronic health conditions, though confidence intervals overlapped (Figure S5). An exception was high blood pressure, where the predicted probability of IN-HLT among those with the condition appeared to be lower in the 1970c than the 1958c. In unadjusted models, having a health condition at age 42 tended to be associated with a lower risk of being ACT-FT at age 50-54. There were no clear associations with ACT-FT, UNEMP, or IN-OTH. Adjusting for economic activity at age 42 (lagged outcome) strongly attenuated associations between health and economic activity, however, associations persisted (Supplementary Material).

In fully adjusted regression models, those with chronic health conditions at age 42 exhibited a higher risk of being IN-HLT in their fifties in both cohorts, accounting for differences in previous economic activity and sociodemographic characteristics between groups (Figure 6). Having a longstanding illness at age 42 was associated with an approximately 6 percentage-point higher risk of being IN-HLT nearly a decade later (AME_1958_ = 5.9 [95% CI 2.7, 9.1], AME_1970_ = 6.5 [95%CI 3.4, 9.6]). Back pain (AME_1958_ = 2.4 [95%CI -0.2, 4.8], AME_1970_ = 3.2 [95%CI 0.3, 5.0]) and psychological distress (AME_1958_ = 4.4 [95%CI 0.9, 7.9], AME_1970_ = 4.5 [95%CI 1.1, 7.8]) were also associated with a higher risk of IN-HLT. There was no strong evidence for an association between obesity and IN-HLT in the 1958c, though there was some suggestion of an association in the 1970c (AME_1970_ = 2.2 [95%CI -0.6, 5.0]). There was some suggestion that having diabetes at age 42 was associated with a higher risk of IN-HLT (AME_1958_ = 4.4 [95% CI -2.1, 10.9], AME_1970_ = 5.3 [95% CI -0.8, 11.4]), but analyses were underpowered, and estimates were imprecise due to the low prevalence of diabetes at age 42 in both cohorts. High blood pressure was associated with a higher risk of being IN-HLT in the 1958c only (AME_1958_ = 4.7 [95%CI 0.6-8.9]). Predicted probabilities of IN-HLT among those without chronic conditions were very similar across cohorts once previous economic activity and socioeconomic factors had been accounted for, as were the predicted probabilities of IN-HLT among those with chronic health conditions (Figure S6). There was some suggestion that those with high blood pressure had a lower predicted probability of IN-HLT in the 1970c, but confidence intervals overlapped. As in the unadjusted models, there was some suggestion that having a health condition at age 42 was associated with a lower risk of being ACT-FT at age 50-54, and there were no clear associations with ACT-FT, UNEMP or IN-OTH.

**Figure 6.**
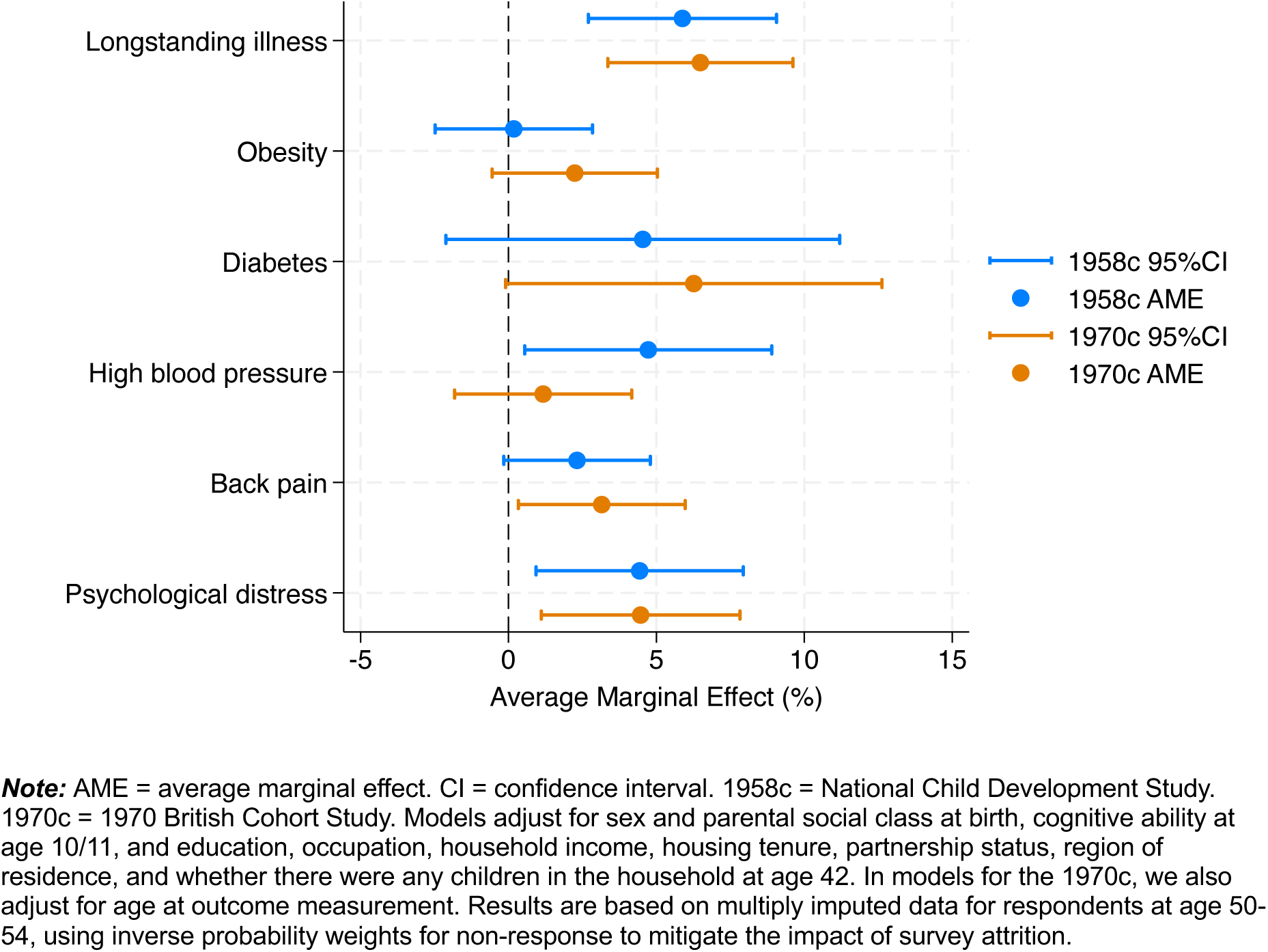
**Average marginal effects from fully adjusted models**.

### Secondary Analysis

We described the gender and socioeconomic composition of those with health conditions at age 42 in the ACT-FT and IN-HLT groups at age 50-54 (Table 2). Within each cohort, those in the ACT-FT group were more likely to be male and socioeconomically advantaged (degree-level qualifications, non-manual occupations, homeowners) while those who in the IN-HLT group were more socioeconomically disadvantaged compared to all those with a chronic condition and to the cohort overall. Though confidence intervals were large, there was some suggestion that the relative disadvantage of those in the IN-HLT group was more pronounced in the 1970c (a higher percentage were in the lowest income quintile and not home owners compared to the 1958c, and similar percentage had no degree-level qualifications and were in manual occupations compared to the 1958c, despite higher education and non-manual occupation levels in the 1970c overall). The proportion of women in the IN-HLT group was also greater in the 1970c, potentially due to higher female labour force participation in the 1970c entitling more women to receive disability benefit and so self-report themselves as IN-HLT.

**Table 2.** Sociodemographic composition of “active full-time” (ACT-FT) and “inactive due to health” (IN-HLT) groups at age 50-54 among those with longstanding illness, obesity, or psychological distress at age 42.

| Health at age 42 | 1958c |  |  |  | 1970c |  |  |  |
| --- | --- | --- | --- | --- | --- | --- | --- | --- |
|  | Has condition | Has condition | Has condition | Any | Has condition | Has condition | Has condition | Any |
| Activity at age 50-54 | ACT-FT | IN-HLT | Any | Any | ACT-FT | IN-HLT | Any | Any |
| **Longstanding illness** |  |  |  |  |  |  |  |  |
| SEX: Male | 62.5<br>(58.8-66.1) | 55.1<br>(47.5-62.7) | 49.7<br>(46.5-53.4) | 50.7<br>(49.2-52.1) | 56.8<br>(52.1-61.5) | 44.1<br>(34.2-54.0) | 44.6<br>(40.8-48.3) | 49.6<br>(47.9-51.3) |
| EDUCATION : No degree | 67.3<br>(63.8-70.7) | 90.0<br>(87.2-92.8) | 74.9<br>(72.6-77.3) | 71.4<br>(70.3-72.5) | 56.7<br>(52.1-61.4) | 87.1<br>(80.5-93.7) | 67.5<br>(64.0-71.0) | 63.1<br>(61.3-64.8) |
| INCOME: Lowest quintile | 17.2<br>(13.4-21.0) | 51.0<br>(41.2-60.7) | 29.1<br>(25.4-32.9) | 21.7<br>(20.2-23.2) | 17.7<br>(13.3-22.1) | 74.2<br>(65.1-83.4) | 36.8<br>(32.4-41.2) | 24.4<br>(22.2-26.7) |
| TENURE: Owns home | 78.1<br>(73.8-82.4) | 40.0<br>(31.4-48.6) | 67.6<br>(64.1-71.1) | 76.7<br>(75.0-78.4) | 73.8<br>(68.8-78.7) | 18.0<br>(9.3-26.6) | 55.9<br>(51.6-60.1) | 67.1<br>(64.8-69.4) |
| OCCUPATION: Manual (among active at age 42) | 38.3<br>(34.0-42.5) | 54.5<br>(35.4-73.7) | 40.3<br>(36.5-44.1) | 41.4<br>(35.9-46.8) | 29.7<br>(25.2-34.3) | 55.3<br>(35.8-74.9) | 32.9<br>(28.3-37.5) | 33.6<br>(30.8-36.4) |
| **Obesity** |  |  |  |  |  |  |  |  |
| SEX: Male | 63.7<br>(58.9-68.4) | 53.1<br>(35.9-70.3) | 51.3<br>(46.9-55.6) | 50.7<br>(49.2-52.1) | 62.1<br>(57.1-67.0) | 38.7<br>(23.5-54.0) | 50.4<br>(45.9-54.09) | 49.6<br>(47.9-51.3) |
| EDUCATION: No degree | 72.2<br>(68.1-76.3) | 88.5<br>(82.3-94.6) | 77.6<br>(74.6-80.7) | 71.4<br>(70.3-72.5) | 62.9<br>(58.6-67.2) | 90.6<br>(81.7-99.5) | 68.6<br>(65.0-72.3) | 63.1<br>(61.3-64.8) |
| INCOME: Lowest quintile | 16.7<br>(12.4-21.0) | 48.7<br>(30.6-66.7) | 24.8<br>(20.7-28.9) | 21.7<br>(20.2-23.2) | 17.8<br>(12.7-22.9) | 71.0<br>(55.5-86.4) | 28.9<br>(24.0-33.8) | 24.4<br>(22.2-26.7) |
| TENURE: Owns home | 78.8<br>(73.6-83.9) | 41.1<br>(24.3-58.0) | 70.9<br>(66.0-75.7) | 76.7<br>(75.0-78.4) | 71.1<br>(66.0-76.3) | 18.0<br>(6.5-29.6) | 61.5<br>(55.7-66.4) | 67.1<br>(64.8-69.4) |
| OCCUPATION: Manual (among active at age 42) | 41.6<br>(35.9-47.4) | 57.8<br>(33.0-82.7) | 43.7<br>(38.6-48.8) | 41.4<br>(35.9-46.8) | 37.4<br>(32.3-42.5) | 57.1<br>(30.3-84.0) | 39.1<br>(33.8-44.4) | 33.6<br>(30.8-36.4) |
| **Psychological distress** |  |  |  |  |  |  |  |  |
| SEX: Male | 54.2<br>(47.4-61.1) | 47.9<br>(36.4-59.4) | 40.3<br>(35.3-45.3) | 50.7<br>(49.2-52.1) | 56.0<br>(49.1-62.9) | 42.8<br>(29.2-56.4) | 43.8<br>(38.7-48.8) | 49.6<br>(47.9-51.3) |
| EDUCATION: No degree | 73.7<br>(68.4-79.0) | 90.0<br>(85.8-93.9) | 80.9<br>(78.0-83.4) | 71.4<br>(70.3-72.5) | 63.3<br>(57.8-68.8) | 89.1<br>(81.4-96.8) | 72.5<br>(68.5-76.5) | 63.1<br>(61.3-64.8) |
| INCOME: Lowest quintile | 21.2<br>(14.0-28.4) | 49.9<br>(36.6-63.2) | 35.0<br>(29.1-40.9) | 21.7<br>(20.2-23.2) | 20.3<br>(13.8-26.8) | 79.4<br>(23.8-35.4) | 41.2<br>(35.6-46.7) | 24.4<br>(22.2-26.7) |
| TENURE: Owns home | 76.7<br>(68.8-84.6) | 31.7<br>(20.1-43.3) | 61.4<br>(56.0-66.6) | 76.7<br>(75.0-78.4) | 68.3<br>(61.7-75.0) | 17.3<br>(7.7-27.0) | 52.8<br>(47.2-58.3) | 67.1<br>(64.8-69.4) |
| OCCUPATION: Manual (among active at age 42) | 39.0<br>(31.5-46.4) | 55.4<br>(31.2-79.6) | 42.3<br>(36.4-48.1) | 41.4<br>(35.9-46.8) | 34.9<br>(28.5-41.4) | 59.1<br>(35.7-82.4) | 37.7<br>(31.7-43.7) | 33.6<br>(30.8-36.4) |

**Note:** Proportions are based on multiply imputed and inverse probability weighted data. ACT-FT = active full-time. IN-HLT = inactive for health reasons. Sex is sex at birth, and education, household equivalised income, housing tenure, and occupation are measured at age 42. The proportion of cohort members in manual occupations is given among those who were economically active at age 42.

### Sensitivity Analysis

Results from sensitivity analyses are presented in the Supplementary Material. Point estimates from fully adjusted models were very similar to those from models that combined ACT-FT, ACT-PT and UNEMP into a single group and when controlling for additional potential early-life confounders. This was also the case when the analytical sample was restricted to those economically active (ACT-FT, ACT-PT or UNEMP) at age 42, and when those IN-HLT at age 42 were excluded.

Having a longstanding illness or psychological distress at age 42 was associated with a higher risk of IN-HLT at age 50-54 in all strata in both cohorts (men vs. women, degree vs. no degree, income quintile 1 (lowest) vs. quintiles 2 to 5, home owner vs. other; Figure S7). Analyses were underpowered to test whether there were differences in associations within strata across cohorts, or within cohorts across strata. Our interpretation therefore focuses on the direction of the effect.

## DISCUSSION

### Interpretation of the Findings

Using rich observational data from two long-running birth cohort studies of people born 12 years apart, we explored the relationship between health at age 42 and economic activity a decade later.

The prevalence of nearly all poor health outcomes was higher in the more recently born cohort (1970c), reflecting a more general trend of “generational health drift” and increasing age-specific prevalence of chronic disease across post-war cohorts born (Gimeno et al., 2024; Gondek et al., 2019; Jivraj et al., 2020). Higher prevalence of poor health in the 1970c was observed for economic activity groups, including those working at age 50-54, which could be indicative of the greater ability of individuals in the 1970c to remain in work despite ill-health (be this due to reasonable adjustments, the changing nature of jobs, or better management of the condition). However, the higher prevalence poor health in the 1970c among the economically active constitutes a larger pool of individuals at risk of health-related inactivity as cohort members grow older. The prevalence of ill-health among those inactive due to health also tended to be higher in the 1970c, and that there was some indication that those inactive due to health problems in the 1970c were more socioeconomically disadvantaged than those in the 1958c.

In both cohorts, chronic health conditions at age 42 were associated with a higher risk of being inactive due to health at age 50-54, consistent with findings from other studies quantifying associations between health and labour market exit in the UK and internationally (Blundell et al., 2023; Britton & French, 2020; Jones et al., 2020; van Rijn et al., 2014).

Evidence was strongest for longstanding illness and symptoms-based conditions (back pain and mental ill-health). Except for high blood pressure, the additional risk of being inactive due to health incurred by those with a health condition compared to their peers was similar across cohorts once previous economic activity and sociodemographic characteristics were controlled for. This is despite changes in social, epidemiologic, and economic contexts experienced by the two cohorts across their lives.

One might expect a weaker association between ill-health and economic inactivity in the 1970c given increasing prevalence of most chronic conditions and a similar proportion of cohort members inactive due to health at age 50-54 across cohorts. However, for most of the conditions included in these analyses, the change in distribution of economic activity across cohorts was similar for those with and without the health conditions.

Accounting for socioeconomic differences and previous economic activity, there was little difference in the predicted probability of being inactive due to health across cohorts for either those with or without chronic health conditions (except for high blood pressure).

There could also be other explanations which future work could seek to disentangle. Multimorbidity has become more common in more recently born cohorts (Gondek, 2020; Ribe et al., 2024). Our analyses did not account for multiple conditions, and it is possible that, for example, those with back pain in the 1970c are also more likely to be experiencing psychological distress. For conditions like high blood pressure and diabetes, other studies have found that associations with labour market exit are driven primarily through comorbidities (Kark et al., 2007; Nexø et al., 2020; Runge et al., 2024). The weaker association between high blood pressure and health-related inactivity in the 1970c could be explained by better diagnosis and treatment, resulting in fewer complications. As well as experiencing more multimorbidity, members of the later born cohort experience the onset of some conditions, such as obesity, at an earlier age (Ervasti et al., 2016; Johnson et al., 2015). There is some evidence that, among individuals reporting the same level of health, a previous history of poor health is associated with a greater likelihood of labour market drop out (Disney et al., 2006), suggesting that both health histories and current health matter for individuals’ labour market decisions. Individuals with chronic health conditions in the 1970c could also perceive their own health as worse. However, examinations of measurement equivalence of the Malaise Inventory (from which the psychological distress is derived) suggest that members of the 1958c and 1970c interpret questionnaire items similarly (Ploubidis et al., 2019).

### Strengths and Limitations

A major strength of this study is its use of data from two large birth cohorts with similar study designs, but representative of people born 12 years apart. We were able to quantify associations between long-term health conditions and subsequent economic activity using the same analytical strategy, enabling us to compare the magnitude of these associations across cohorts. With data collection spanning over fifty years, we were able to examine the impact of health on economic activity nearly a decade later, which is important considering this study’s focus on chronic conditions, the effects of which likely play out across long time horizons. Since data was collected prospectively, self-reported health at age 42 could not be affected by economic activity at age 50-54. The rich data collected in the British birth cohorts on health, social, and economic factors enabled us to adjust for a wide range of potential confounders measured during both childhood and adulthood, including cognitive ability, as well as previous economic activity. This strengthens the plausibility of the assumption of no unmeasured confounding – although it cannot be completely ruled out – which is crucial for interpreting the findings as indicative of causal effects.

This study also has several limitations. Long-term health conditions were self-reported and operationalised as a binary variable, so our measure does not capture the severity of these conditions (whether due to the condition itself or to comorbidities). We assumed that awareness of having a health condition (either due to experiencing symptoms or receiving a diagnosis), a pre-requisite for self-reporting that condition, is an important part of the mechanism linking poor health and labour market exit, especially for chronic health conditions which often do not wholly limit the ability to work (O’Donnell et al., 2015). New cases will also have arisen between age 42 and 50-54, which could result in the impact of poor health on work participation being underestimated since these individuals would be included in the “healthy” comparator group.

Additionally, some retrospective harmonisation was needed to produce comparable indicators of health conditions across cohorts. This mean that for some conditions, we had to use “ever” rather than “current” measures at age 42. For conditions like back pain, which can be intermittent, use of “ever” measures, the risk of health-related inactivity may be underestimated since the exposure measure will also include those who suffered from back pain in the past but have recovered. Obesity, psychological distress and longstanding illness are “current” measures, and cohort members may transition in and out of these states between ages 42 and 50-54, potentially resulting in risk of health-related inactivity being underestimated for these health conditions. Nevertheless, we see clear associations between mental ill-health and back pain with health-related inactivity, potentially linked to the difficulty in seeking treatment for and managing recurrent health conditions.

Another limitation is the potential impact of period effects on exposure and outcome measurement. Measurement of economic activity at age 50-51 in the 1958c coincided with the start of the 2008 Great Recession. Some studies have shown that the likelihood of claiming disability benefit increases during times of financial crisis, since rates of job loss are often higher among those with health problems, and disability benefits are often more generous and have lower conditionality than unemployment benefits (Pasini & Zantomio, 2013). Future research could seek to expand analyses to other birth cohorts and age combinations or to use the full employment histories available for 1958c and 1970c cohort members to overcome this (Hancock, 2016; Hancock & Peters, 2021).

Finally, like all longitudinal studies, 1958c and 1970c experienced attrition over time. We attempted to mitigate bias from loss to follow-up through a combination of multiple imputation and inverse probability weighting. However, loss to follow-up did result in a smaller analytical cohort. Despite >7000 respondents at age 50-54 in each cohort, analyses were underpowered because of the low prevalence of some health conditions (e.g., diabetes) age 42, and small numbers of individuals transitioning between activity states. This particularly impacted our ability to explore whether associations differed for population subgroups. While we could show a consistent direction in the association between health and health-related economic inactivity, we could not draw conclusions on whether the magnitude of associations differed across strata within cohorts, or across cohorts within strata. Other studies using larger survey datasets and administrative data have suggested that there is a difference in the magnitude of associations between men and women, and by educational attainment (Jones et al., 2020; Trevisan & Zantomio, 2016). Future work may seek to pool birth cohort data.

### Implications

Chronic health conditions in early midlife are strongly associated with health-related inactivity a decade later, in cohort members’ early fifties, a decade before State Pension Age. Transitions into economic activity only make up part of the overall economic cost of chronic health conditions. Among people who are economically active, poor health is associated with reductions in working hours (Jones et al., 2020), and increased sickness absence (Bryan et al., 2021). Considering the impact of health on labour supply, productivity, lost tax revenue, and increased benefit expenditure and health and care costs, the OECD has estimated that the economic cost of overweight/obesity and mental-ill health is approximately 4% of UK Gross Domestic Product, respectively (OECD, 2019; OECD & European Union, 2018).

Improving population health through increased prevention and a focus on the wider determinants of health is important not only in its own right, but also for the economy (Crawshaw et al., 2024; Scott, 2021a, 2021b). In the UK, transitioning back into economic activity for those receiving long-term disability benefits remains rare (Latimer et al., 2024). Only 1-in-6 people return to economic activity withing the first year of becoming inactive due to health problems, and this rapidly declines to 1-in-20 for those who have been out of work for a year or more (Office for Budget Responsibility, 2023). It is therefore also crucial to better support people with chronic conditions to remain or return to economic activity, for instance by improving the management of chronic conditions once they are diagnosed (including through occupational health) and considering the impact of mental-physical multimorbidity (Henderson et al., 2024).

## Funding Sources

This research was funded by the Medical Research Council (grant number MR/N013867/1 to LG) and the European Research Council (grant number ERC-2021-CoG-101002587 to JBD). The Leverhulme Trust (grant number RC-2001-003) supports the Leverhulme Centre for Demographic Science at the University of Oxford. The Economic and Social Research Council (grant numbers ES/M001660/1 and ES/W013142/1) supports the Centre for Longitudinal Studies at University College London. The funders had no role in the design of execution of this study, nor in the decision to publish.

## Ethics

Ethical approval for the National Child Development Study (NCDS) and the 1970 British Cohort Study (BCS70) was granted by the NHS Multi-Centre Research Ethics Committee. All participants gave informed consent.

## Declaration of Interest

The authors declare no competing interests.

## Data Statement

All data used in this study can be accessed by registered users free of charge through the UK Data Service (https://ukdataservice.ac.uk).

## Author Contributions

Laura Gimeno: conceptualisation, data curation, formal analysis, investigation, methodology, visualisation, writing – original draft, writing – review and editing. Charis Bridger Staatz: conceptualisation, data curation, methodology, writing – review and editing. Alice Goisis: conceptualisation, supervision, writing – review and editing. Jennifer B. Dowd: conceptualisation, supervision, writing – review and editing. George B. Ploubidis: conceptualisation, funding acquisition, methodology, supervision, writing – review and editing.

## Data Availability

All data used in this study can be accessed by registered users free of charge through the UK Data Service.

https://ukdataservice.ac.uk

## Notes

### Competing Interest Statement

The authors have declared no competing interest.

