## Supplement A for "Chronic health conditions and health-related economic inactivity in midlife: Evidence from the 1958 and 1970 British birth cohorts"

**Table S1.** Description of chronic health condition exposure measures used at age 42 in the 1958c and 1970c.

| | NCDS | BCS70 | Analysis variable |
|---|---|---|---|
| Longstanding illness | Do you have any longstanding illness, disability or infirmity that has troubled you over a period of time or is likely to affect you over a period of time? | Do you have any physical or mental health conditions or illnesses lasting or expected to last 12 months or more? | 0 = No longstanding illness<br>1 = Longstanding illness |
| Psychological distress[1] | 9-Item Malaise Inventory, using a cut-off score of ≥4 (range 0-9). | 9-Item Malaise Inventory, using a cut-off score of ≥4 (range 0-9). | 0 = No psychological distress<br>1 = Psychological distress |
| Obesity[2] | Body mass index ≥30 kg/$m^2$ from harmonised self-reported height and weight. | Body mass index ≥30 kg/$m^2$ from harmonised self-reported height and weight. | 0 = No obesity<br>1 = Obesity |
| Diabetes[3] | Harmonised indicator of lifetime diabetes prevalence, leveraging all parent reported and self-reported information from birth. | Harmonised indicator of lifetime diabetes prevalence, leveraging all parent reported and self-reported information from birth. | 0 = Never diabetes<br>1 = Ever diabetes |
| High blood pressure | Lifetime prevalence indicator from ever report of high blood pressure at age 42. | Lifetime prevalence indicator derived from ever report of back pain at age 30, and reports of back pain since last sweep at ages 34, 38 and 42. | 0 = Never high blood pressure<br>1 = Ever high blood pressure |
| Back pain | Lifetime prevalence indicator from ever report of back pain at age 42. Question asks about whether cohort member has experienced persistent back pain, sciatica, or slipped disc. | Lifetime prevalence indicator derived from ever report of back pain at age 30, and reports of back pain since last sweep at ages 34, 38 and 42. Questions ask about whether cohort member has experienced persistent back pain, sciatica, or slipped disc. | 0 = Never back pain<br>1 = Ever back pain |

[1]This measure has been shown to exhibit scalar invariance across the two cohorts, suggesting that members of both cohorts interpret the items of the Malaise Inventory similarly (Ploubidis, Sullivan, Brown and Goodman, 2017. *Psychol Med;* 47(2): 291-303).

[2]Hardy, Johnson and Park, 2016. CLOSER Work Package 1: Harmonised height. Weight and BMI User Guide. London: CLOSER.

[3]Gimeno, Narayanan and Hardy, 2025. Harmonised indicators of self-reported diabetes in five British birth cohort studies. User Guide (Version 1). London: Centre for Longitudinal Studies.

**Table S2.** Prevalence of chronic health conditions at age 42.

| | 1958c<br>% (95% CI) | 1970c<br>% (95% CI) |
|---|---|---|
| Longstanding illness | 30.8 (29.8-31.8) | 31.1 (29.9-32.3) |
| Obesity | 16.8 (15.9-17.7) | 23.9 (22.6-25.2) |
| Diabetes | 2.6 (2.3-3.0) | 3.3 (2.9-3.8) |
| High blood pressure | 11.9 (11.2-12.6) | 13.5 (12.7-14.3) |
| Back pain | 23.1 (22.2-24.1) | 31.0 (29.9-32.1) |
| Psychological distress | 14.2 (13.4-15.0) | 21.4 (20.1-22.6) |

***Note:*** 1958c = National Child Development Study. 1970c = 1970 British Cohort Study. Analysis was carried out on imputed and inverse-probability weighted data to account for nonresponse.

**Table S3.** Distribution of the 1958c and 1970c by economic activity at ages 42 and 50-54.

| | 1958c<br>% (95% CI) | 1970c<br>% (95% CI) |
|---|---|---|
| **Age 42** | | |
| ACTFT | 64.8 (63.7–65.8) | 63.3 (62.1–64.6) |
| ACTPT | 17.6 (16.9–18.4) | 18.4 (17.4–19.3) |
| UNEMP | 2.7 (2.3–3.1) | 3.6 (3.0–4.2) |
| INHLT | 5.8 (5.2–6.4) | 5.0 (4.4–5.6) |
| INOTH | 9.1 (8.5–9.8) | 9.7 (8.9–10.5) |
| **Age 50-54** | | |
| ACTFT | 64.0 (62.6–65.4) | 64.6 (62.8–66.4) |
| ACTPT | 15.9 (15.0–17.0) | 16.8 (15.6–18.1) |
| UNEMP | 3.3 (2.7–3.9) | 2.0 (1.4–2.7) |
| INHLT | 8.5 (7.5–9.4) | 8.8 (7.4–10.2) |
| INOTH | 8.2 (7.3–9.2) | 7.7 (6.8–8.7) |

***Note:*** 1958c = National Child Development Study. 1970c = 1970 British Cohort Study. Analysis was carried out on imputed and inverse-probability weighted data to account for nonresponse.

**Table S4.** Distribution of economic activity at age 42 within categories of economic activity at age 50-51 in 1958c (n = 9761).

| | | **Activity @50** | | | | |
|---|---|---|---|---|---|---|
| | | ACTFT | ACTPT | UNEMP | INHLT | INOTH |
| **Activity @42** | ACTFT | 85.9 | 21.7 | 53.9 | 32.2 | 20.6 |
| | ACTPT | 8.0 | 59.9 | 11.0 | 10.5 | 17.8 |
| | UNEMP | 2.0 | 1.3 | 27.4 | 3.2 | 0.7 |
| | INHLT | 1.2 | 1.5 | 1.4 | 46.7 | 1.4 |
| | INOTH | 2.9 | 15.6 | 6.3 | 7.5 | 59.5 |

***Note:*** 1958c = National Child Development Study. ACTFT = active full-time. ACTPT = active part-time. UNEMP = unemployed. INHLT = inactive due to health reasons. INOTH = inactive due to other reasons. Analysis was carried out on imputed and inverse-probability weighted data to account for nonresponse.

**Table S5.** Distribution of economic activity at age 42 within categories of economic activity at age 51-54 in the 1970c (n = 7336).

| | | Activity @51-54 | | | | |
|---|---|---|---|---|---|---|
| | | ACTFT % | ACTPT % | UNEMP % | INHLT % | INOTH % |
| **Activity @42** | ACTFT | 84.3 | 22.5 | 28.9 | 22.3 | 31.8 |
| | ACTPT | 10.0 | 60.2 | 16.9 | 14.3 | 14.5 |
| | UNEMP | 2.1 | 2.1 | 45.2 | 6.9 | 5.1 |
| | INHLT | 0.4 | 0.6 | 1.9 | 50.2 | 0.8 |
| | INOTH | 3.2 | 14.5 | 7.0 | 6.2 | 47.8 |

**Table S6.** Prevalence of poor health at age 42 by economic activity at age 50-54.

| | **Economic activity at age 50-54** | | | | |
|---|---|---|---|---|---|
| **Health at 42** | **ACTFT % (95%CI)** | **ACTPT % (95%CI)** | **UNEMP % (95%CI)** | **INHLT % (95%CI)** | **INOTH % (95%CI)** |
| **1958c** | | | | | |
| LSI | 25.8 (23.7-27.8) | 27.4 (23.9-30.9) | 33.2 (19.8-46.6) | 72.8 (64.8-80.8) | 37.1 (30.0-44.2) |
| Obesity | 17.1 (15.2-19.0) | 12.4 (9.4-15.4) | 20.0 (8.1-31.9) | 21.4 (14.9-27.8) | 25.5 (17.6-33.5) |
| Diabetes | 2.1 (1.4-2.7) | 2.9 (1.2-4.6) | 3.2 (0.1-7.8) | 5.1 (1.9-8.4) | 2.8 (0.1-5.5) |
| HIBP | 10.3 (8.8-11.8) | 11.9 (9.2-14.6) | 17.4 (7.6-27.2) | 24.1 (16.2-32.0) | 16.8 (10.4-23.2) |
| Back Pain | 21.5 (19.7-23.3) | 21.3 (18.2-24.5) | 36.1 (24.2-48.0) | 38.6 (30.8-46.3) | 26.5 (19.8-33.1) |
| PD | 9.9 (8.5-11.2) | 14.7 (11.6-17.9) | 27.4 (14.8-40.1) | 38.3 (30.2-46.5) | 25.6 (18.1-33.0) |
| **1970c** | | | | | |
| LSI | 23.8 (21.8-25.9) | 30.7 (25.5-35.9) | 36.6 (15.6-57.6) | 80.5 (72.0-89.0) | 36.9 (29.3-44.5) |
| Obesity | 24.5 (22.3-26.8) | 16.4 (12.9-19.9) | 18.1 (2.5-33.8) | 37.8 (27.3-48.6) | 22.1 (15.7-28.6) |
| Diabetes | 2.9 (2.3-3.5) | 2.6 (1.1-4.0) | 1.0 (0.1-2.4) | 9.5 (4.6-14.5) | 3.6 (1.6-5.6) |
| HIBP | 12.6 (11.4-13.9) | 10.9 (8.5-13.3) | 15.5 (2.2-28.8) | 20.7 (13.0-28.3) | 17.1 (12.2-22.0) |
| Back Pain | 29.3 (27.4-31.2) | 32.8 (28.5-37.1) | 30.0 (13.1-47.0) | 47.0 (36.9-57.2) | 31.7 (25.1-38.3) |
| PD | 16.7 (14.7-18.8) | 18.9 (14.8-23.1) | 29.0 (10.2-47.8) | 56.9 (45.6-68.1) | 25.1 (16.6-33.6) |

***Note:*** ACTFT = active full-time. ACTPT = active part-time. UNEMP = unemployed. INHLT = inactive due to health reasons. INOTH = inactive due to other reasons. CI = confidence interval. 1958c = National Child Development Study. 1970c = 1970 British Cohort Study. LSI = longstanding illness. HIBP = high blood pressure. PD = psychological distress. Analysis was carried out on imputed among respondents at age 50-54 and inverse-probability weighted data to account for nonresponse.

**Table S7.** Demographic and socioeconomic characteristics by cohort among respondents to age 50/51-54 sweeps.

| | 1958c (n = 9761) % (95% CI) | 1970c (n = 7337) % (95% CI) |
|---|---|---|
| **Sex at birth** | | |
| Male | 50.7 (49.2-52.1) | 49.6 (47.9-51.3) |
| Female | 49.3 (47.9-50.8) | 50.4 (48.7-52.1) |
| **Education at age 42** | | |
| Degree (NVQ 4-5) | 28.6 (27.5-29.7) | 36.9 (35.2-38.7) |
| No degree (NVQ 0-3) | 71.4 (70.3-72.5) | 63.1 (61.3-64.8) |
| **Occupation at age 42** | | |
| Manual (RGSC IIIM-V) | 41.4 (35.9-46.8) | 33.6 (30.8-36.4) |
| Non-manual (RGSC I-IIINM) | 58.6 (53.2-64.1) | 66.4 (63.4-69.1) |
| **Equivalised household income at age 42** | | |
| Lowest quintile | 21.7 (20.2-23.2) | 24.4 (22.2-26.7) |
| 2 | 20.4 (20.0-21.8) | 20.5 (18.6-22.4) |
| 3 | 19.4 (18.0-20.8) | 19.5 (17.9-21.1) |
| 4 | 19.7 (18.4-21.1) | 18.3 (16.8-19.7) |
| Highest quintile | 18.8 (17.4-20.2) | 17.3 (16.0-18.9) |
| **Housing tenure at age 42** | | |
| Owns or is buying own home | 76.7 (75.0-78.4) | 67.1 (64.8-69.4) |
| Other | 23.3 (21.6-25.0) | 32.9 (30.6-35.2) |
| **Partnership status at age 42** | | |
| Married or cohabiting | 78.2 (76.4-80.0) | 59.0 (56.8-61.3) |
| Not married or cohabiting | 21.8 (20.2-23.6) | 41.0 (38.7-43.2) |
| **Parental social class at age 42** | | |
| Partly/unskilled (RGSC IV-V) | 22.3 (20.7-23.9) | 22.5 (20.7-24.3) |
| Managerial/intermediate/skilled (RGSC I-III) | 77.7 (76.1-79.3) | 77.5 (75.6-79.6) |
| **Any dependent children at age 42** | | |
| Yes | 73.8 (72.1-75.5) | 71.8 (70.0-73.8) |
| No | 26.2 (24.5-27.9) | 28.2 (26.2-30.1) |
| **Region of residence[a] at age 42** | | |
| North of England | 24.9 (23.1-26.8) | 26.9 (24.9-28.9) |
| West of England | 19.3 (17.5-21.1) | 19.1 (17.4-20.9) |
| East of England | 43.0 (41.1-44.9) | 42.0 (40.0-44.1) |
| Wales/Scotland/Northern Ireland | 12.8 (11.8-13.8) | 11.9 (10.6-13.2) |

***Note:*** Means and percentage shown in the table are based on imputed data weighted for non-response. Except sex at birth, all other variables were measured at age 42. Patterns of missing data are shown in Table S2. RGSC = Registrar General Social Class 1990. NVQ = National Vocational Qualification. [a]North of England = North East, North West, Yorkshire & The Humber. West of England = West Midlands, South West. East of England = East Midlands, East Anglia, South East and London.

**Figure S1.** Timeline of major disability legislation in the United Kingdom since 1970.

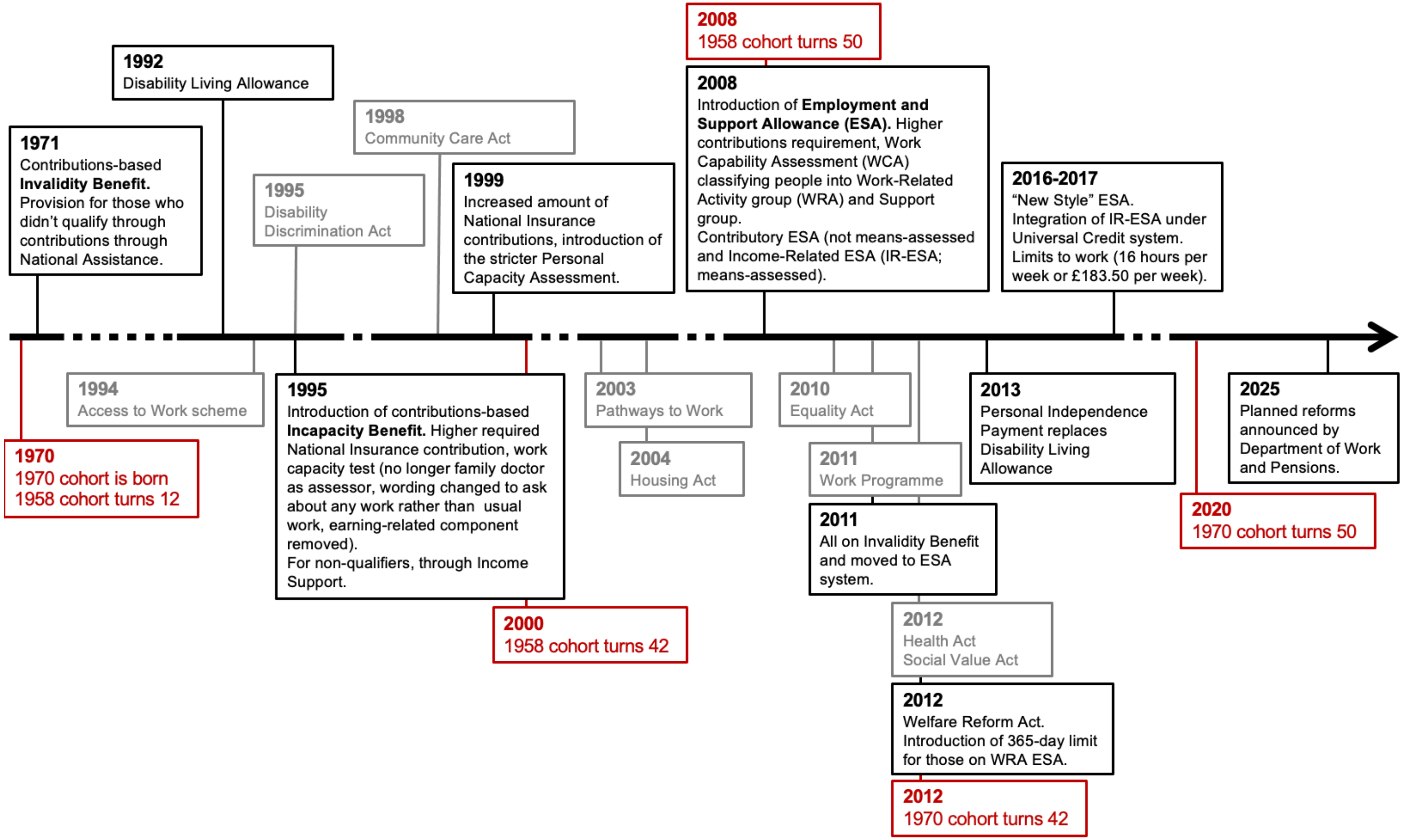

***Note:*** Legislation and reforms related to the disability benefits system are shown in black. Legislation protecting the rights of those with disabilities, anti-discrimination laws and policies aiming to encourage participation of those with disability in employment are shown in grey. Major life events for the 1958 and 1970 cohorts are shown in red.

**Figure S2.** Percentage of cohort members who were unemployed, economically inactive due to poor health, or economically inactive due to other reasons in the period surrounding the start of the 2008 Financial Crisis in the United Kingdom (January 2008).

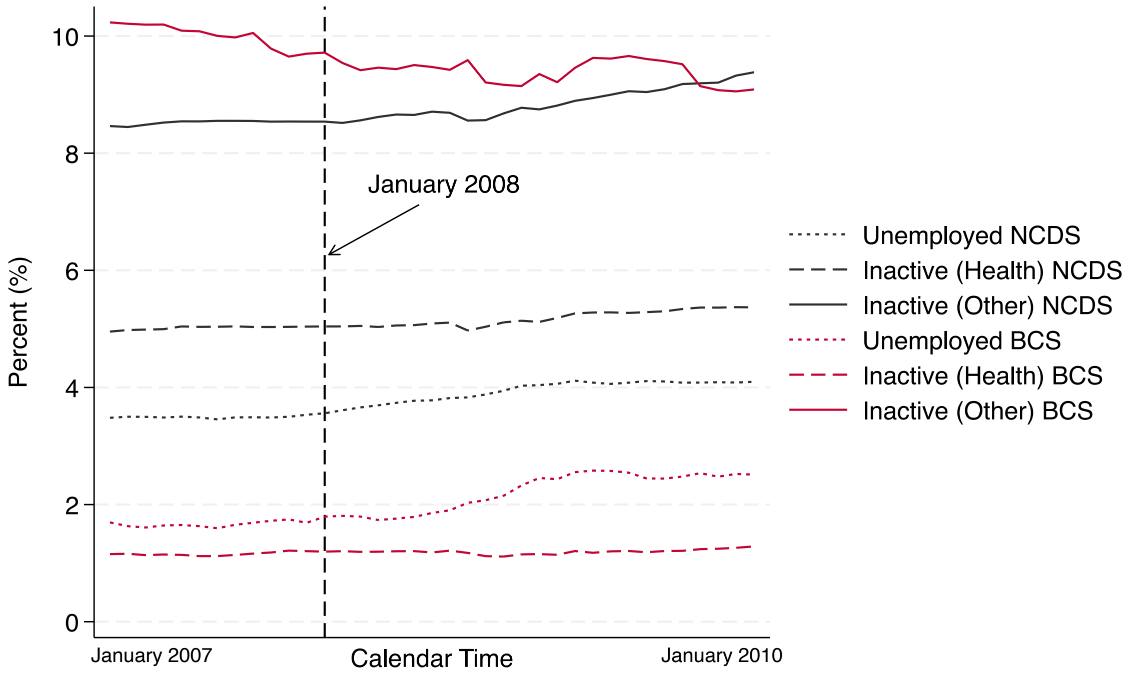

***Note:*** This graph show inactivity and unemployment rates for the 1958c and 1970c between January 2007 and January 2010, using monthly activity history data (Hancock & Peters, 2021; Hancock, 2016). Since the x-axis is calendar time, members of the two cohorts were different ages in this same time window (1958c aged 49-52 and 1970c aged 37-40), which explains the higher rate of health-related inactivity in the 1958c. Though there was an upward trend in unemployment in both cohorts, this was more marked in the 1970c cohort, while in the 1958c individuals may have been more likely to transition into inactivity.

**Figure S3.** Prevalence of chronic health conditions at age 42 within economic activity groups at age 50-54 in the 1958c and 1970c.

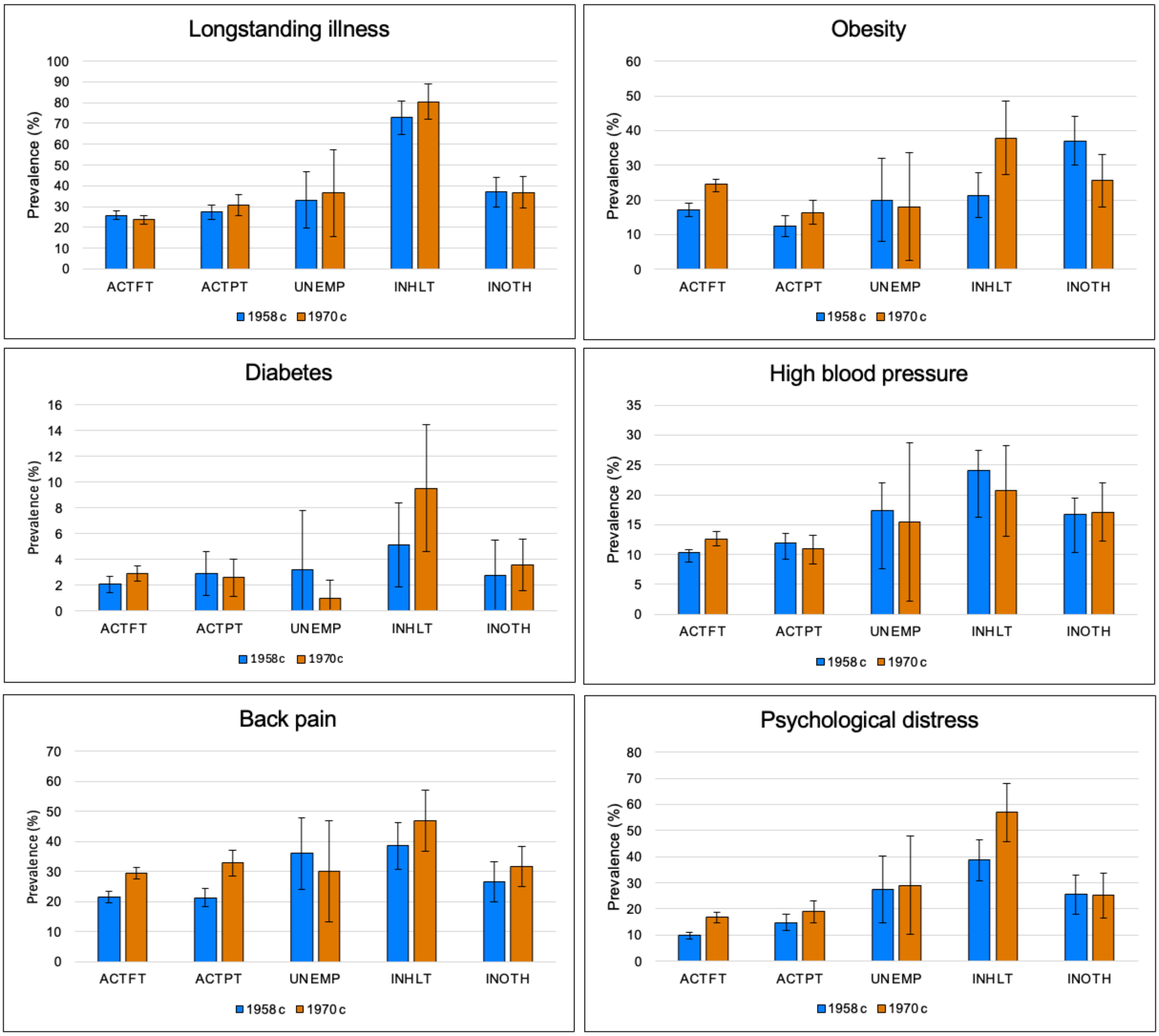

***Note:*** ACTFT = active full-time. ACTPT = active part-time. UNEMP = unemployed. INHLT = inactive due to health reasons. INOTH = inactive due to other reasons. 1958c = National Child Development Study. 1970c = 1970 British Cohort Study. Underlying values are given in Table S4.

**Figure S4.** Distribution of individuals with and without chronic health conditions at age 42 by economic activity at age 50-54 in the 1958c and 1970c.

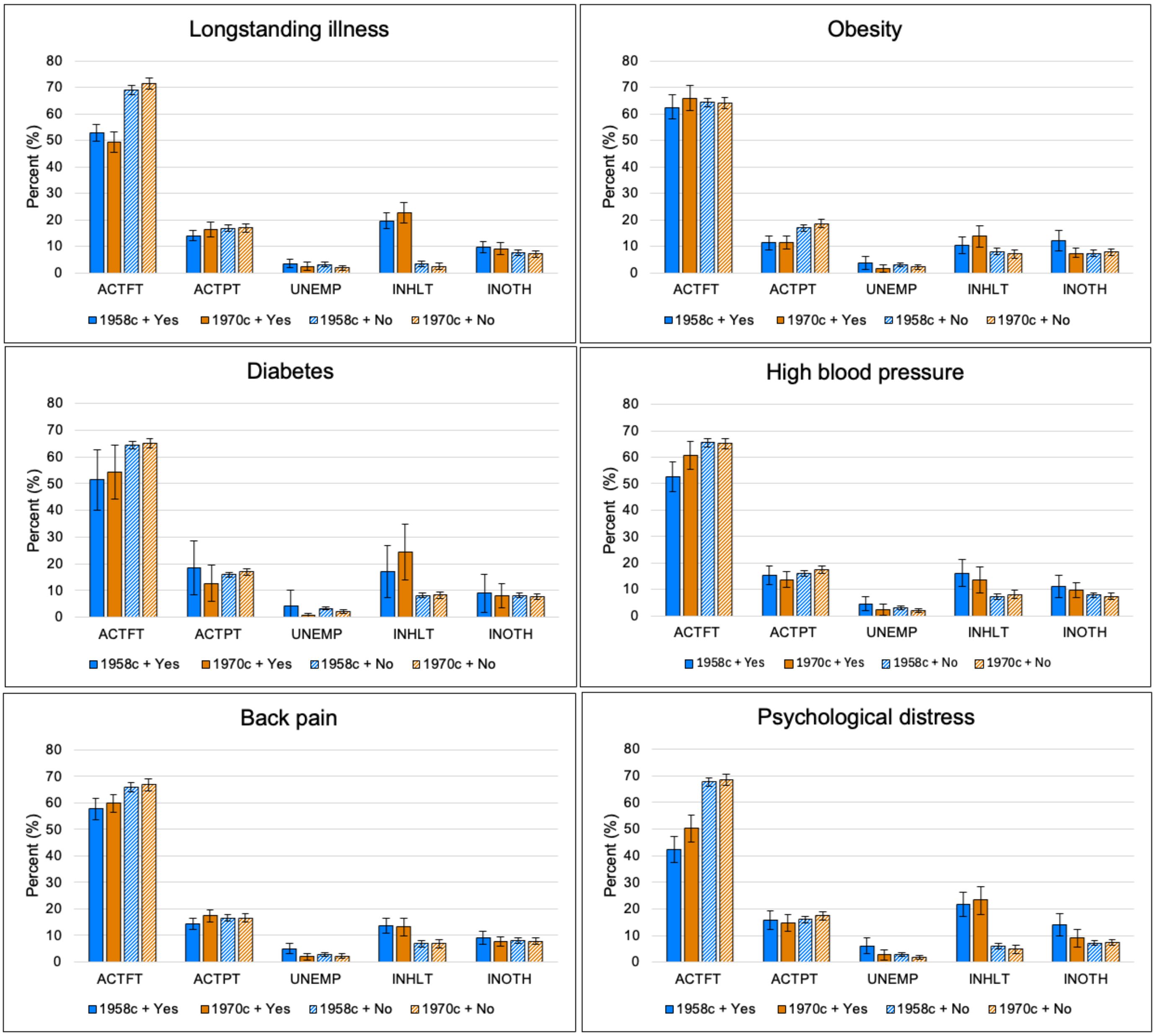

***Note:*** ACTFT = active full-time. ACTPT = active part-time. UNEMP = unemployed. INHLT = inactive due to health reasons. INOTH = inactive due to other reasons. 1958c = National Child Development Study. 1970c = 1970 British Cohort Study. Yes = Among those with the health condition. No = Among those without the health condition.

**Figure S5.** Predicted probability of being inactive due to health problems (INHLT) at age 50-54 for those with and without and with chronic health problems in the 1958c and 1970c (Unadjusted).

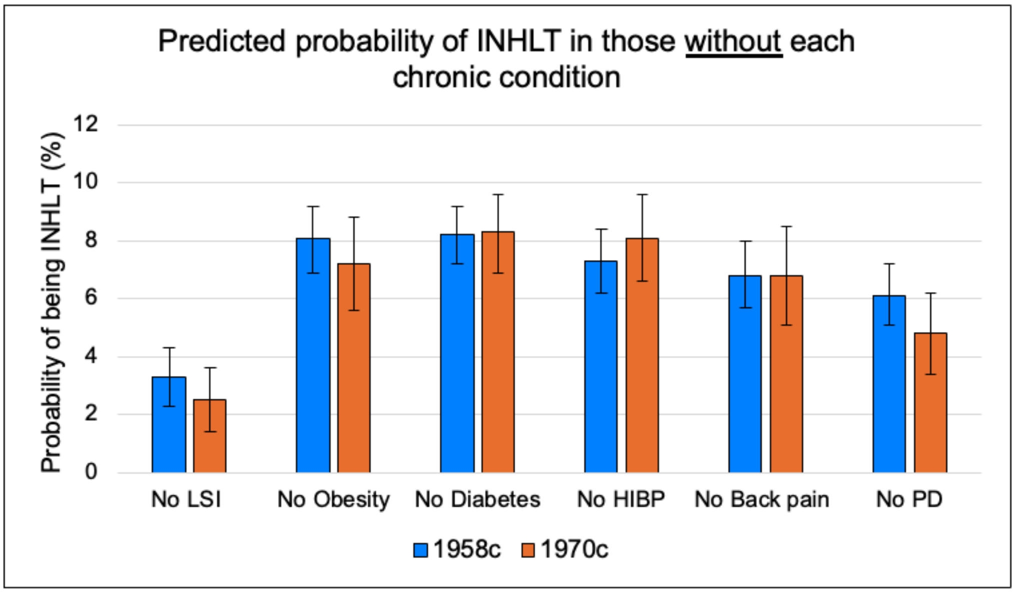

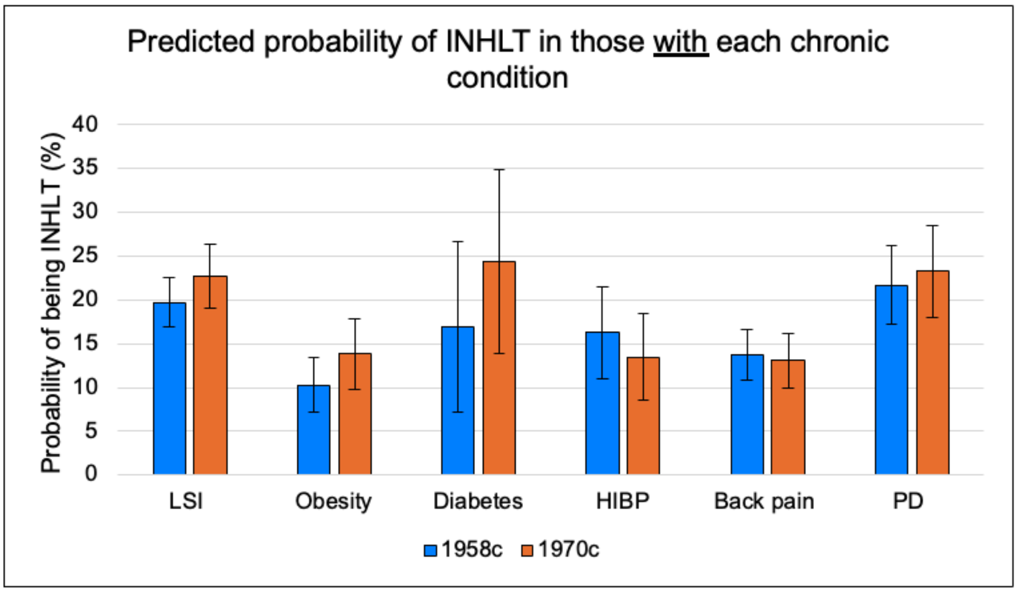

***Note:*** 1958c = National Child Development Study. 1970c = 1970 British Cohort Study. INHLT = health-related economic inactivity. LSI = longstanding illness. HIBP = high blood pressure. PD = psychological distress. Models include each exposure in turn and the outcome variable (economic activity at age 50-54). Here we show the predicted probability of being inactive due to health problems (INHLT) for individuals without each chronic condition. Average Marginal Effects presented in the main manuscript quantify the percentage-point increase in risk of INHLT for those who have the condition of interest relative to this baseline probability. All analyses were conducted on multiply imputed and inverse probability-weighted data to account for item and unit non-response.

**Figure S6.** Predicted probability of being inactive due to health problems (INHLT) at age 50-54 for those with and without chronic health problems in the 1958c and 1970c accounting for previous economic activity and sociodemographic characteristics (Fully adjusted).

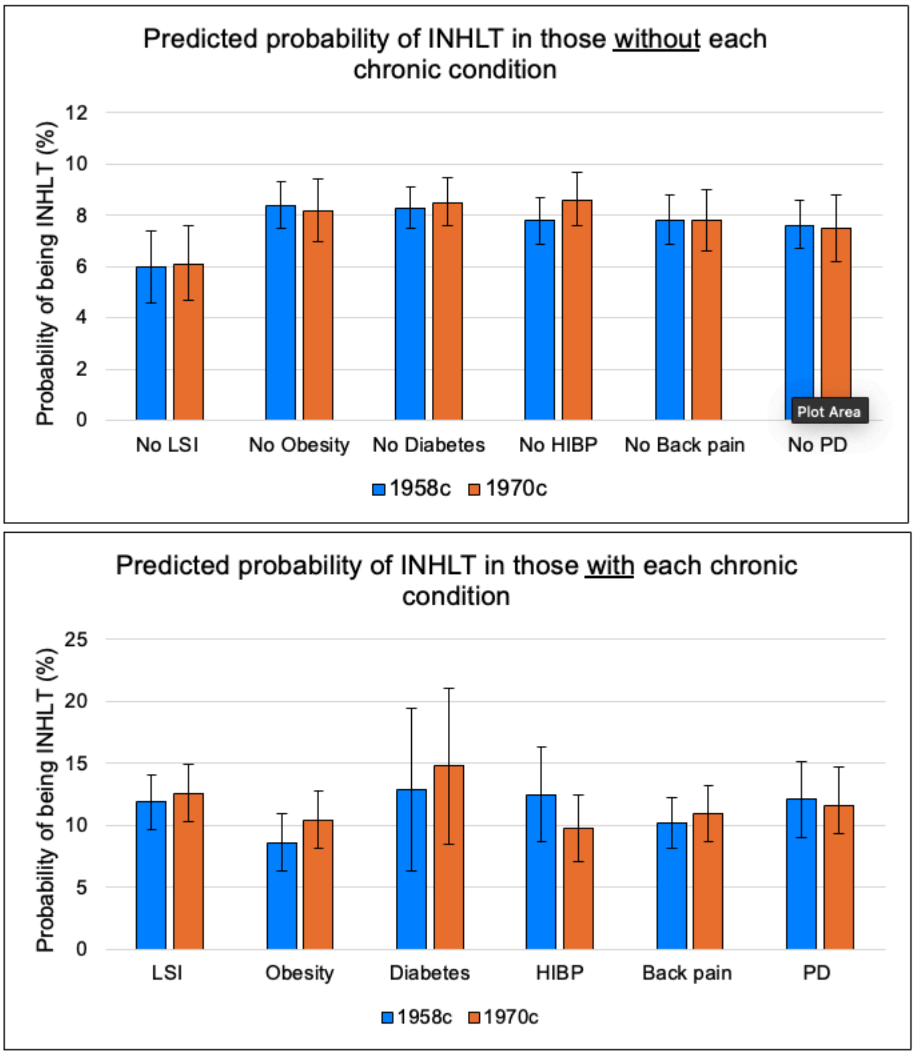

***Note:*** 1958c = National Child Development Study. 1970c = 1970 British Cohort Study. LSI = longstanding illness. HIBP = high blood pressure. PD = psychological distress. Models are adjusted for sex and parental social class at birth, cognitive ability at age 10/11, and education, occupation, household income, housing tenure, partnership status, region of residence, and whether the cohort member had any children in the household at age 42. In models for the 1970c, we also adjust for age at outcome measurement. Here we show the predicted probability of being inactive due to health problems (INHLT) for individuals without each chronic condition. Average Marginal Effects presented in the main manuscript quantify the percentage-point increase in risk of INHLT for those who have the condition of interest relative to this baseline probability. All analyses were conducted on multiply imputed and inverse probability-weighted data to account for item and unit non-response.

**Figure S7.** Average marginal effects from fully adjusted, stratified models for longstanding illness and psychological distress.

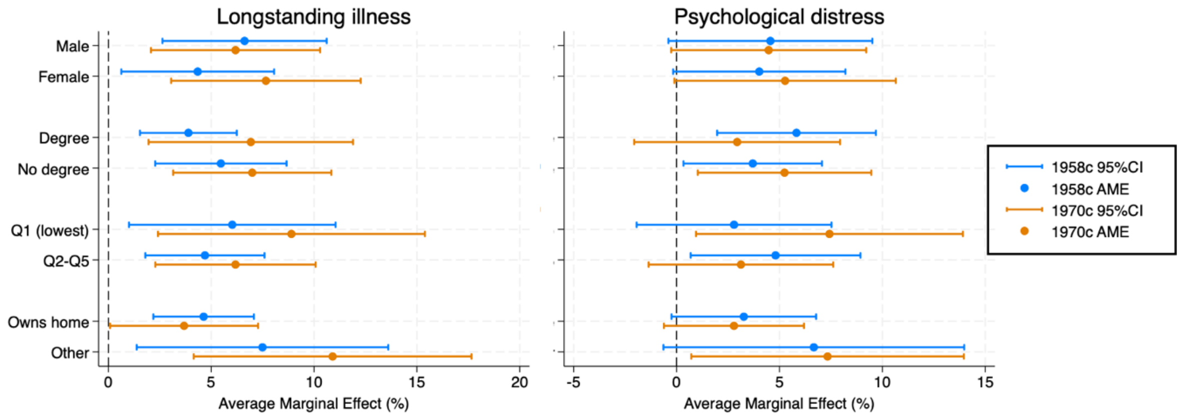

***Note:*** AME = average marginal effect. CI = confidence interval. 1958c = National Child Development Study. 1970c = 1970 British Cohort Study. Models are adjusted for sex and parental social class at birth, cognitive ability at age 10/11, and education, occupation, household income, housing tenure, partnership status, region of residence, and whether the cohort member had any children in the household at age 42. In models for the 1970c, we also adjust for age at outcome measurement. The outcome variable is categorical with three levels: active, inactive due to health, and inactive due to other reasons (with active as the baseline group). All analyses were conducted on multiply imputed and inverse probability-weighted data to account for item and unit non-response.

Note that analyses were underpowered to test whether there were differences in associations within strata across cohorts, or within cohort across strata. As such, we focus our interpretation on the direction of the effect across strata.

**Text S1.** Missing data strategy.

Like all longitudinal studies, the 1958c and 1970c experience loss to follow-up, the likelihood of which varies by individual level characteristics, such as gender, health, and socioeconomic status. Over time, this results in the respondents being a non-representative subset of the cohort as it was initially sampled. We capitalised on the richness of the cohort data and the known properties of sample to restore sample representativeness at age 42 and 50-54, combining multiple imputation to handle item non-response, and inverse probability weights (IPW) to handle unit non-response. We used a similar approach for both the generation of descriptive statistics (chronic health condition prevalence, distribution by economic activity) at age 42, and to create the analysis datasets from which regression models were run.

To deal with item missingness amongst respondents at age 42 and/or at age 50-54, we imputed missing data using multivariate imputation by chained equations (MICE). Multiple imputation works under the assumption that data are Missing At Random (MAR), that is, that the observed data can explain systematic differences between the observed and missing values (White, Royston & Wood, 2011). However, this assumption is largely untestable. To make the MAR assumption more plausible, we included data on exposures, the outcome, all covariates, and a diverse set of auxiliary variables predictive of non-response or of the underlying missing values shown in Table S8 below (Mostafa et al, 2021; Katsoulis et al, 2024). Information on the amount of missing data is shown Table S9 below.

To account for unit non-response at age 42 and/or at age 50-54 (that is, people not responding to the survey sweep of interest), we derived IPWs for non-response. Weights were derived for the target population at each sweep, which is cohort members who were alive and living in the UK at the point of data collection (e.g., at age 50-54). Multiple imputation was used to create the data from which inverse probability weights were derived, to make sure that all cohort members could be included in the weight derivation process. We imputed data on all variables to be used in the regression models (one imputation model per sweep per study), creating 5 imputed datasets for each cohort. Each imputation model contained an indicator of sweep non-response (the outcome), and a range of predictors of non-response measured earlier in the lifecourse. These included sex at birth, parental social class, household overcrowding in childhood, cognitive ability and mental health in childhood/adolescence, measures of social capital/social participation (e.g., voting, membership in social/political organisations, social support, partnership status), socioeconomic status (e.g., educational attainment, whether in employment, income), health in adulthood (e.g., mental health, body mass index, self-rated general health, smoking status), and previous response to sweeps (i.e., number of previous sweeps cohort member had responded to).

Following imputation, we constructed logistic regression models (one per cohort) to predict the probability of responding to each sweep using imputed predictors (associated with non-response. We used the regression output (predicted probability of responding to the sweep based on observed characteristics) to create inverse probability weights. We truncated weights to the value of 10 to prevent extreme weights exerting undue influence on our analyses. The weights were then rescaled to the respective sweeps, so that the sum of the weights was equal to the number of respondents at the sweep. The weighted distribution of sex at birth and parental socioeconomic status at birth in the age 42 and 50-54 analytical samples were similar to the distribution of these characteristics in the birth sweep.

**Table S8.** Variables used in multiple imputation for the descriptive analyses at age 42 (among respondents at age 42; Model 1) and for the main analysis (among respondents at age 50-54; Model 2).

| Variable | Description | Age measured | | In model | |
|---|---|---|---|---|---|
| | | 1958c | 1970c | 1 | 2 |
| ***Exposures*** | | 42 | 42 | | |
| LSI | Yes or no (binary) | 42 | 42 | X | X |
| PD | Yes or no (binary) | 42 | 42 | X | X |
| Obesity | Yes or no (binary) | 42 | 42 | X | X |
| Back pain | Ever or never (binary) | 42 | 42 | X | X |
| Diabetes | Ever or never (binary) | 42 | 42 | X | X |
| HIBP | Ever or never (binary) | 42 | 42 | X | X |
| ***Outcome/Lagged Outcome*** | | | | | |
| Economic activity 50-54 | ACTFT, ACTPT, UNEMP, INHLT, INOTH (categorical) | 50-51 | 51-54 | | X |
| Economic activity 42 | ACTFT, ACTPT, UNEMP, INHLT, INOTH (categorical) | 42 | 42 | X | X |
| ***Socioeconomic controls or auxiliary variables*** | | | | | |
| Mother's education | Age left full-time education (continuous) | 0 | 0 | X | X |
| Father's education | Age left full-time education (continuous) | 0 | 0 | X | X |
| Father's social class | Professional/managerial/intermediate, or partly skilled/unskilled | | | X | X |
| Housing tenure childhood | Parents owned home or other (binary) | 7 | 5 | X | X |
| People per room | People in household divided by number of rooms in house excluding kitchen and bathrooms (ordinal/continuous) | 7 | 5 | X | X |
| Previous housing tenure | Owned home or other (binary) | 33 | 34 | X | X |
| Previous economic activity | ACTFT, ACTPT, UNEMP, INHLT, INOTH (categorical) | 33 | 34 | X | |
| Previous employment | Working or not working (binary) | 33 | 34 | | X |
| Previous household income | Quintiles of equivalised income (ordinal) | 33 | 34 | X | X |
| Previous education | Degree (NVQ 4-5) or no degree (NVQ 0-3) (binary) | 33 | 34 | X | X |
| Previous manual occupation | Manual (RGSC IIIM-V) or non-manual (RGSC I-IIINM) (binary) | 33 | 34 | X | X |
| Housing tenure | Owned home or other (binary) | 42 | 42 | | X |
| Household income | Quintiles of equivalised income (ordinal) | 42 | 42 | | X |
| Education | Degree (NVQ 4-5) or no degree (NVQ 0-3) (binary) | 42 | 42 | | X |
| Manual occupation | Manual (RGSC IIIM-V) or non-manual (RGSC I-IIINM) (binary) | 42 | 42 | | X |
| ***Demographic controls or auxiliary variables*** | | | | | |
| Sex at birth | Male or female | 0 | 0 | X | X |
| Country of birth | England or other (binary) | 0 | 0 | X | X |

| | | | | | |
|---|---|---|---|---|---|
| Number of children | Number of dependent children (continuous) | 33 | 34 | X | X |
| Previous partnership status | Married/cohabiting or other (binary) | 33 | 34 | X | X |
| Whether has children | Whether has ≥1 dependent child (binary) | 42 | 42 | | X |
| Partnership status | Married/cohabiting or other (binary) | 42 | 42 | | X |
| Region of residence | North of England, West of England, East of England, Wales/Scotland/Northern Ireland (categorical) | 42 | 42 | | X |
| Age at outcome measurement | Age at interview in years (continuous) | NA | 51-54 | | X |
| ***Health controls or auxiliary variables*** | | | | | |
| Birthweight | Birthweight in grams (continuous) | 0 | 0 | X | X |
| Whether breastfed | Ever or never (binary) | 0 | 0 | X | X |
| Mother smoked pregnant | Yes or no (binary) | 0 | 0 | X | X |
| Cognitive ability | Principal component 1 (continuous) | 11 | 10 | X | X |
| Child medical conditions | Whether had any of five conditions in the last year: pathological heart condition, recurrent sore throat, recurrent abdominal pain, eczema, hay fever (continuous) | 11 | 10 | X | X |
| Child mental health | Rutter total score (continuous) | 11 | 10 | X | X |
| Child BMI | Body mass index in kg/m$^2$ (continuous) | 11 | 10 | X | X |
| Adolescent mental health | Malaise Inventory total score (continuous) | 16 | 16 | X | X |
| Adolescent BMI | Body mass index in kg/m$^2$ (continuous) | 16 | 16 | X | X |
| Maternal mental health | Malaise Inventory total score (continuous) | NA | 16 | X | X |
| Previous adult mental health | Malaise Inventory total score (continuous) | 33 | 34 | X | X |
| Previous adult BMI | Body mass index in kg/m2 (continuous) | 33 | 34 | X | X |
| Previous adult LSI | Yes or no (binary) | 33 | 34 | X | X |
| Smoking | Current or non-smoker (binary) | 33 | 34 | X | X |
| Self-reated health | Excellent-Good or Fair-Poor (binary) | 33 | 34 | X | X |
| ***Auxiliary variable: Previous response patterns*** | | | | | |
| Number of previous sweeps | Number of sweeps participated in prior to age 50/51-54 sweep (continuous) | 0-33 OR 0-46 | 0-38 OR 0-46 | X | X |

***Note:*** LSI = longstanding illness. PD = psychological distress. BMI = body mass index. HIBP = high blood pressure. NVQ = National Vocational Qualification. RGSC = Registrar General Social Class.

**Table S9.** Number and percentage of observations among respondents at age 50-54 with missing information on exposures and covariates.

| | NCDS (N = 9761) n (%) | BCS70 (N = 7336) n (%) |
|---|---|---|
| **Exposures** | | |
| Longstanding illness at age 42 | 697 (7.1) | 598 (8.2) |
| Obesity at age 42 | 1565 (16.0) | 1788 (24.4) |
| Diabetes at age 42 | 696 (7.1) | 25 (0.3) |
| High blood pressure at age 42 | 703 (7.2) | 236 (3.2) |
| Back pain at age 42 | 700 (7.2) | 234 (3.2) |
| Psychological distress at age 42 | 768 (7.9) | 1261 (17.2) |
| **Covariates** | | |
| Sex at birth | 0 (0) | 0 (0) |
| Education at age 42 | 1 (<0.1) | 573 (7.8) |
| Occupation at age 42 | 1952 (20.0) | 1448 (19.7) |
| Household income quintile at age 42 | 2059 (21.1) | 1821 (24.8) |
| Housing tenure at age 42 | 727 (7.5) | 601 (8.2) |
| Partnership status at age 42 | 724 (7.4) | 575 (7.8) |
| Cognitive ability at age 10/11 | 1327 (13.6) | 2031 (27.7) |
| Region of residence at age 42 | 690 (7.1) | 570 (7.8) |
| Children in household at age 42 | 704 (7.2) | 570 (7.8) |
| Parental occupation at birth | 932 (9.6) | 568 (7.7) |
| Economic activity at age 42 | 695 (7.1) | 582 (7.9) |
